# Leveraging large-scale genetics of PTSD and cardiovascular disease demonstrates robust shared risk and improves risk prediction accuracy

**DOI:** 10.1101/2022.07.25.22278004

**Authors:** Antonia V. Seligowski, Burook Misganaw, Lucie A. Duffy, Kerry J. Ressler, Guia Guffanti

## Abstract

Individuals with posttraumatic stress disorder (PTSD) are significantly more likely to be diagnosed with cardiovascular disease (CVD; e.g., myocardial infarction, stroke). The evidence for this link is so compelling that the National Institutes of Health convened a working group to determine gaps in the current literature, including the need for large-scale genomic studies to identify shared genetic risk. The current study used a large healthcare biobank dataset of *N* = 36,412 individuals, combined with GWAS summary statistics from publicly available, large-scale PTSD and CVD studies. We found significant genetic correlations between PTSD and CVD (r_G_=0.24, S.E.= 0.05982, *p=5*.*09E-05*), and Mendelian randomization analyses indicated a potential causal link from PTSD to hypertension (*β*=0.20, S.E.= 0.04, *p=5*.*97e-6*), but not the reverse. PTSD summary statistics significantly predicted PTSD diagnostic status (*R*^*2*^=0.27%, *p*=*5*.*31E-08*), and this was significantly improved by incorporating summary statistics from CVD and major depressive disorder (*R*^*2*^=1.30%, *p*=*2*.*16E-32*). Further, pathway enrichment analyses indicated that genetic variants involved in shared PTSD-CVD risk included those involved in postsynaptic structure (*p=0*.*00001*), synapse organization (*p=0*.*0001*), and interleukin-7 (IL-7) mediated signaling pathways (*p=0*.*0002*). While additional research is needed to determine the clinical utility of these findings, these data further support the biological shared risk for stress-related disorders of mental health and cardiovascular illness.

## Introduction

Cardiovascular disease (CVD) is the leading cause of death in the United States, affecting over 650,000 people each year.^1^ Rates of CVD and hypertension are even higher among those who have experienced trauma, and particularly those with posttraumatic stress disorder (PTSD).^2,3^ PTSD is a highly impairing psychiatric condition with symptoms such as intrusions, avoidance of trauma-related stimuli, negative alterations in cognition and mood, and hyperarousal.^4^ PTSD is often characterized by an exaggerated fear response controlled by the sympathetic nervous system. It is thought that chronic activation of this system strains the cardiovascular and immune systems, ultimately increasing susceptibility to CVD.^5^ Despite the epidemiological evidence linking PTSD and CVD, there remain significant gaps with regard to how PTSD and CVD interact to confer risk in trauma-exposed individuals.

Given the preponderance of evidence supporting a PTSD-CVD link, the National Heart, Lung, and Blood Institute (NHLBI) convened a working group entitled, “*The Cardiovascular Consequences of Post-traumatic Stress Disorder,”* which brought together experts from the NHLBI and the American Heart Association to determine research gaps and priorities.^6^ As reviewed by O’Donnell et al. (2021),^7^ there are several gaps related to the genetics of PTSD and CVD: 1) *“Lack of large-scale datasets with deep genetic data and both PTSD and CVD measures;”* 2) *“Lack of population cohort studies and biobanks to harness genetics and genomics to understand shared risks of PTSD and CVD;”* and 3) *“Lack of polygenic risk scores from genome-wide association studies (GWAS) of PTSD and CVD to inform potential causal PTSD-CVD associations and identify potential therapeutic targets*.*”* Addressing these gaps will improve PTSD and CVD risk identification, which can be used to improve PTSD treatment and reduce concurrent CVD risk. While there is still much to be done to address these gaps, the PTSD and CVD genetics fields have made significant advances with the advent of new technologies and more international collaboration.

Recent PTSD genetic studies have primarily taken a GWAS approach catapulted by the creation of the PGC-PTSD (the PTSD division of the Psychiatric Genomics Consortium).^8^ The first GWAS analyses in PTSD identified several risk genes and single nucleotide polymorphisms (SNPs), including the *rs8042149* SNP,^9^ the Tolloid-Like 1 gene,^10^ *lincRNA AC068718*.*1*,^11^ the PRTFDC1 gene,^12^ and SNP *rs717947*.^8,13^ In the first PGC-PTSD study, Duncan et al.^14^ found significant genetic overlap between PTSD and schizophrenia, and recent work identified sex- and ancestry-based differences in PTSD heritability^15^. The PTSD GWAS summary statistics from the PGC-PTSD have also been used to create polygenic risk scores (PRS) that predict PTSD in large independent cohorts of Veterans.^15,16^ Given the heterogeneity of PTSD and its polygenic nature, PRS studies using PGC-PTSD summary statistics will be crucial to improving our prediction of PTSD risk.

The field of CVD genetics has seen similar advances in terms of utilizing large samples across research groups. The Coronary ARtery DIsease Genome wide Replication and Meta-analysis (CARDIoGRAM) consortium is comparable to the PGC-PTSD in that it was developed to allow for significantly larger studies with more power to detect genetic contributions to coronary artery disease (CAD; the largest subset of CVD).^17^ The consortium initially conducted a meta-analysis of GWASs and identified 13 risk loci for CAD, and several follow-up studies confirmed most of these and found evidence for additional loci implicating genes involved in vascular function (e.g., *PECAM1, PROCR*).^18-20^ GWAS has also been used to identify genetic risk for other aspects of CVD and its risk factors, such as blood pressure,^21^ stroke,^22^ atrial fibrillation,^23^ carotid intima media thickness and carotid plaque (i.e., subclinical atherosclerosis),^24^ and heart failure.^25^

Three studies have reported on the shared genetics of PTSD and CVD. Using a candidate gene approach, Pollard and colleagues^26^ found that 37 PTSD risk genes relevant to immune function and inflammation were also independent risk genes for CVD. Sumner et al.^27^ used the PGC-PTSD GWAS summary statistics^12^ and found evidence for genetic overlap of PTSD and CAD, but these results did not survive multiple testing. In a subsequent study, Sumner et al.^28^ used PGC-PTSD GWAS summary statistics and found significant main effects of PTSD symptoms for blood pressure in a trans-ethnic meta-analysis, although there was heterogeneity in the results. Given these promising findings, additional large-scale genetic analyses are needed to better account for the shared heritability of PTSD and CVD, and to improve genetic prediction. Further, it is critical to consider major depressive disorder (MDD) because it is highly comorbid with PTSD,^29^ and like PTSD, MDD carries a significant risk of CVD.^30^ Thus, genetic risk studies will be enhanced by considering PTSD, MDD, and CVD as inherently linked disorders.

The current study addressed gaps in the PTSD-CVD genetics literature as identified by the NHLBI working group.^6^ Specifically, we utilized a large-scale healthcare biobank dataset of *N* = 36,412 individuals that contains CVD, PTSD, and MDD diagnostic data, as well as both PTSD and CVD GWAS summary statistics that included 1.3 million samples. Our goals were to (i) test genetic correlations between PTSD/MDD and CVD, and estimate their magnitudes using current GWAS summary statistics data, (ii) leverage these relationships to improve genetic prediction performances, and (iii) identify the shared biological pathways/genes underlying the genetic correlations.

## Methods

### Participants and procedure

This study included *N* = 36,412 participants from the Mass General Brigham Biobank (MGBB), which is a biorepository of Mass General Brigham Healthcare (e.g., Massachusetts General Hospital, McLean Hospital). Participants provide blood samples, complete health surveys, and consent to these being linked to clinical data from Electronic Health Records (EHR). This study included the subset of MGBB subjects who had their blood samples genotyped as of April 2020. PTSD diagnosis was determined based on the presence of at least one ICD-10 code in a patient’s EHR (i.e., based on clinical judgment). Hypertension and depression phenotypes were determined using Curated Disease Populations determined using algorithms developed by the MGBB to increase phenotype accuracy (see Supplemental material).^31,32^ Cardiovascular disease sub-phenotypes of congestive heart failure and ischemic stroke were also determined through Curated Disease Populations while myocardial infarction was determined based on the presence of at least one ICD-10 code. Pulse and average systolic and diastolic blood pressure were also collected from EHR through the MGBB. Averages were determined from all of the measures of these vital signs available in individual health histories. Therefore, the number of collections included in the average differs for each patient. See Table 1 for more information.

**Table 1:** Descriptive statistics for MGBB sample

|  | PTSD |  | No PTSD |  | Overall |  |
| --- | --- | --- | --- | --- | --- | --- |
|  | *N* | % | *N* | % | *N* | % |
| **Gender** |  |  |  |  |  |  |
| Female | 1459 | 67.90% | 18253 | 53.30% | 19712 | 54.10% |
| Male | 689 | 32.10% | 16009 | 46.70% | 16698 | 45.90% |
| **Predicted ancestry** |  |  |  |  |  |  |
| AFR | 267 | 12.40% | 2144 | 6.30% | 2411 | 6.60% |
| EAS | 21 | 1.00% | 525 | 1.50% | 546 | 1.50% |
| EUR | 1658 | 77.20% | 30092 | 87.80% | 31750 | 87.20% |
| SAS | 202 | 9.40% | 1501 | 4.40% | 1703 | 4.70% |
| **MDD** |  |  |  |  |  |  |
| Cases | 1133 | 52.70% | 5034 | 14.70% | 6167 | 16.90% |
| Controls | 1015 | 47.30% | 29228 | 85.30% | 30243 | 83.10% |
| **CAD** |  |  |  |  |  |  |
| Cases | 121 | 5.60% | 3604 | 10.50% | 3725 | 10.20% |
| Controls | 2027 | 94.40% | 30658 | 89.50% | 32685 | 89.80% |
| **Hypertension** |  |  |  |  |  |  |
| Cases | 910 | 42.40% | 15198 | 44.40% | 16108 | 44.20% |
| Controls | 1238 | 57.60% | 19064 | 55.60% | 20302 | 55.80% |
|  | *Mean* | *SD* | *Mean* | *SD* | *Mean* | *SD* |
| **Age** | 50.5 | 15.3 | 60.4 | 17 | 59.8 | 17.1 |
| **Pulse** | 81.4 | 10.5 | 75.7 | 10.7 | 76 | 10.7 |
| **Systolic BP** | 123 | 11.1 | 127 | 12.4 | 126 | 12.4 |
| **Diastolic BP** | 75.3 | 7.07 | 75.1 | 7.32 | 75.1 | 7.31 |

*Note.* AFR = African; EAS = East Asian; EUR = European; SAS = South Asian; MDD = major depressive disorder; CAD = coronary artery disease; CHF = coronary heart failure; MI = myocardial infarction; SD = standard deviation; BP = blood pressure.

### Genetic data

See Figure 1 for an overview of the genetic analysis procedure. Briefly, de-identified genetic data was requested from the MGBB and genotyped using Illumina SNP array. QC was performed on each batch individually and merged together, resulting in a total of 36,422 individuals and 9,036,179 variants after imputation. Ancestry prediction was performed using a classification model trained on 1000-genome samples.^39^ Most participants included in the summary statistics are of EUR ancestry (>87%). To minimize the effect of population stratification, only those genetically predicted to have EUR ancestry (*n*=31,634) were included for further analysis. Self-identified and predicted ancestry sample sizes are shown in Table S1.

**Figure 1:**
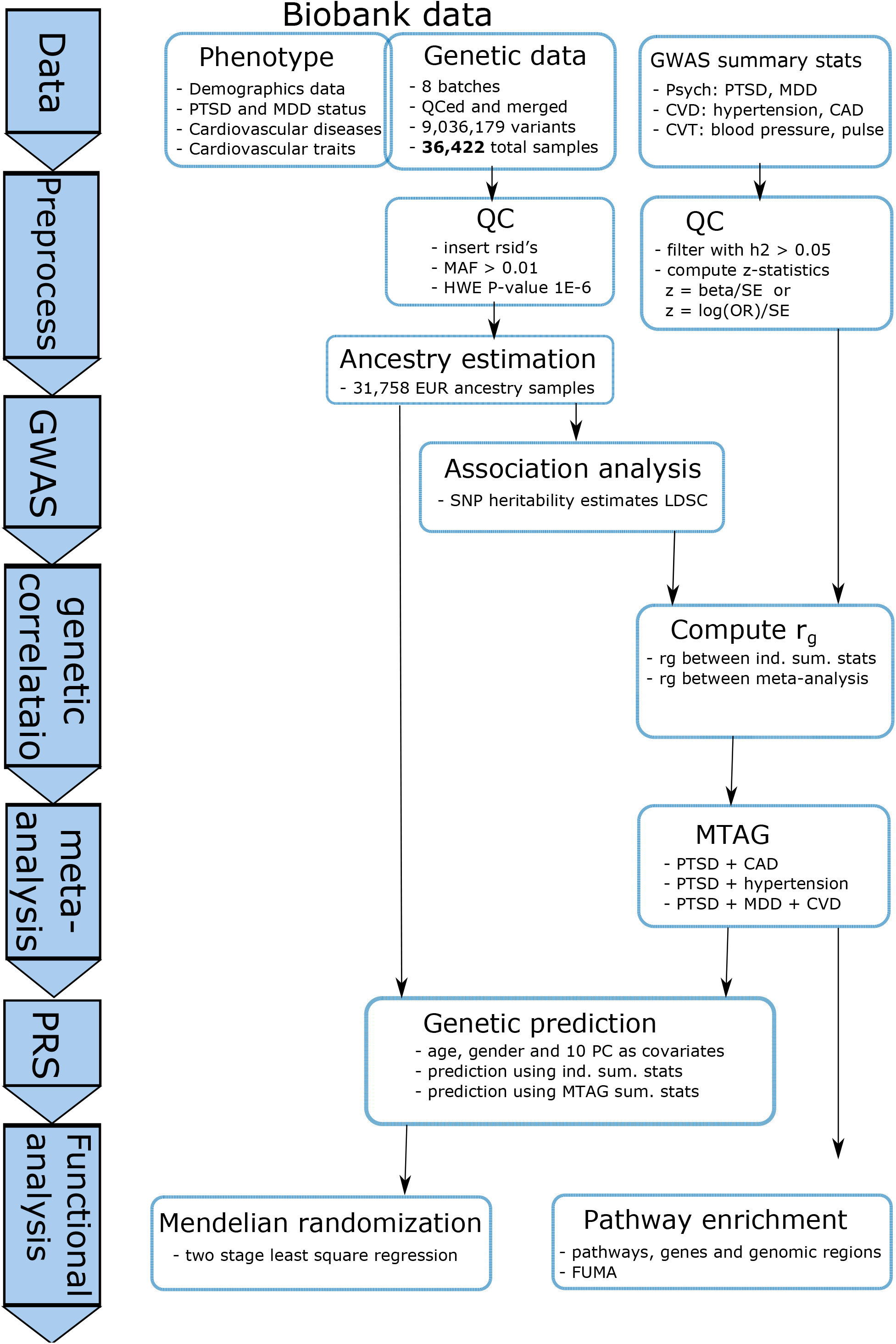
Workflow of data acquisition, processing, and analysis.

### GWAS summary statistics data

Publicly available GWAS summary statistics from several recent publications on PTSD, MDD, and CVD were collected. Summary statistics datasets were processed with the following QC criteria: (i) SNP heritability h^2^ > 0.05, (ii) compute z-statistics (z=beta/SE or z=log(OR)/SE), (iii) filter non-variable SNPs (0<freq<1), and (iv) filter SNPs with low number of samples. Five summary statistics survived the QC criteria and were used for further analysis herein: (i) ptsd_eur_freeze2: the PGC-PTSD freeze-2,^15^ (ii) PGC_MDD2018_ex23andMe: the latest result from PGC-MDD excluding the 23andme samples,^33^ (iii) UKBB_broad_Depression: UKBB analysis for depression phenotypes,^34^ and large-scale genetic studies for (iv) UKBB_Essential_Hypertension: hypertension^35^ and (v) CAD_Nikpay: CAD.^19^ More detail on summary statistics can be found in Table S2.

For proprietary reasons, the PGC-MDD summary statistics that included the 23andme samples are not publicly available. Therefore, we only used summary statistics conducted without the 23andme cohort. With UKBB, summary statistics on three depression phenotypes were provided: probable MDD, ICD-defined MDD, and broad depression (defined by self-reported help-seeking history for problems related to anxiety and depression).^34^ Only broad depression resulted in SNP heritability estimate of greater than 5%. Therefore, only broad depression summary statistics were included for further analysis.

### GWAS and SNP heritability estimates on MGBB data

We conducted a GWAS for four diseases: PTSD, MDD, hypertension, and CAD. A quantitative GWAS was also conducted for quantitative cardiovascular traits (average/minimum/maximum pulse rate, systolic and diastolic blood pressure). Approximately 6M variants were tested. Only variants on the 22 autosomal chromosomes were considered. To control for population stratification, the first 10 principal components were included as covariates. Age, sex, and batch were included as additional covariates. The analysis was performed using PLINK-2.^36^

### Combining summary statistics and estimating genetic correlations

The multi-trait analysis of GWAS (MTAG) approach was used to combine GWAS summary statistics.^37^ MTAG is a generalization of inverse-variance weighted meta-analyses of summary statistics. However, unlike standard meta-analysis, MTAG computes trait-specific association analysis for each individual trait. Summary statistics for PTSD, MDD, hypertension, and CAD were incorporated.

LD score regression (LDSC) was used to estimate genetic correlations.^38^ LDSC is a Python-based command line tool for estimating genetic correlations and SNP heritability from GWAS summary statistics. Precomputed LD scores for EUR ancestry from the 1000 Genomes Project were used.^39^ Genetic correlations were computed among individual summary statistics as well as among meta-analysis summary statistics for the four phenotypes (PTSD, MDD, hypertension, and CAD).

### Constructing and evaluating polygenic risk score

PRS was computed as a weighted sum of the additively coded number of risk/effect alleles (i.e., 0 for homozygous non-risk genotype, 1 for heterozygous genotype, and 2 for homozygous risk genotype). The weights were the beta or odds-ratio from univariate test statistics obtained from the summary statistics of previous individual GWAS or with combined summary statistics using MTAG. Owing to methodological advantage (particularly, not requiring individual level training genetic data) LD clumping followed by P-value thresholding (C+T) is the popular way of selecting variants included in the score. The LD clumping step was done on a window of 250 kb and squared correlation of allele counts of 0.1. In the P-value thresholding step, PRS was computed for several thresholds. The optimal threshold is determined based on prediction performance as assessed by Nagelkerke R^2^ (a psudo-R^2^ for binary variables). Nagelkerke R^2^ was computed as the difference of its value in the full model (which contains the PRS and covariates) and in the null model (which contains only the covariates). This is done for each of the PRS independently. See Figure S1 for results of the thresholding step. PRS analysis was conducted using PRSice-2,^40^ including age, sex, and the first ten ancestry-related principal components as covariates.

To determine the prediction power of PRS, additional performance metrics were evaluated. Area under the ROC curve (AUC) was computed with the *R* statistical software package pROC.^41^ Binary disease labels along with quantitative PRS values were used. To further evaluate the discriminatory ability of MTAG-PRS, stratification performance of the PRS was computed. This was done by binning/grouping samples into equal-sized risk strata/groups based on PRS values. Then, disease incidence rate and odds-ratio were computed for each risk group.

For odds-ratio, the first (the least risk group) was used as reference (odds were computed relative to that group).

### Mendelian randomization

Mendelian randomization was used to test the causal link between PTSD and CVD (see Supplementary Material for more information). Using two stage least square regression (2SLS), PTSD and MDD were our two exposure variables and hypertension and CAD were the two outcome variables, and the same models were tested in reverse with hypertension/CAD as the exposure variables and PTSD/MDD as the outcome variables (i.e., eight models tested). In the first stage, the exposure (binary PTSD or MDD label) is regressed against PTSD/MDD PRS and control variables (age and the first five principal components). Using this regression model, predicted value of the exposure is computed. In the second stage, the outcome variable (binary hypertension or CAD label) is regressed against the predicted exposure variable and control variables. Then, (epidemiological) causality is inferred based on the significance of the coefficient of the predicted exposure variable.

### Pathway analysis

Pathway analysis was performed on FUMA.^42^ FUMA is a web-based application that annotates a set of given variants from GWAS summary statistics by utilizing information from 18 data repositories and tools. It was implemented in two main analysis steps. In the first step (SNP2GENE), after taking a set of input variants, it added variants that were in linkage disequilibrium with the input significant variants, SNP’s were annotated (with biological functionality) and mapped to genes (with positional, eQTL and chromatin interaction mapping). In the second step (GENE2FUNC), the set of prioritized genes from the first step were annotated with biological functions and mechanisms. This included enrichment in biological pathways and functional categories. Adjusted P-values with Benjamini-Hochberg FDR (False Discovery Rate) for multiple test correction method is computed.

## Results

In the total sample, 2,149 participants were PTSD-positive cases and 34,269 were PTSD-negative controls. In terms of depression, 6,167 were predicted to have current or past history of MDD (of these 1,133 had comorbid PTSD), while the remaining 30,243 were MDD negative. See Table 1 for additional phenotypes and quantitative data by PTSD status.

### Genetic correlations between PTSD and CVD

Genetic correlations were estimated in two ways. First, estimates were calculated using individual GWAS summary statistics (both publicly available as well as from new MGBB GWAS analysis). Then, large-scale meta-analysis summary statistics were computed by combining publicly available as well as the MGBB summary statistics for each of the four disease labels of primary interest. The four meta-analyses were: PTSD (*n* = 183,000): MGBB GWAS + PGC-PTSD freeze-2, MDD (*n* = 362,584): MGBB GWAS + PGC-MDD + UKBB_ICD_MDD, Hypertension (*n* = 440,089): MGBB GWAS + UKBB_Essential_Hypertension, and CAD (*n* = 217,000): MGBB GWAS + Nikpay_CAD. Using both of these approaches, statistically significant genetic correlations were estimated between PTSD and hypertension/CAD. With the meta-analysis summary statistics, PTSD demonstrated a significant positive genetic correlation with hypertension (r_G_=0.35, S.E.=0.06, *p=9*.*45E-10*) and CAD (r_G_=0.24, S.E.= 0.06, *p=5*.*09E-05*). Similarly, MDD demonstrated significant genetic correlations with hypertension (r_G_=0.31, S.E.= 0.03, *p=4*.*73E-33*) as well as CAD (r_G_=0.18, S.E.= 0.03, *p=8*.*99E-09*). Genetic correlation estimates between PTSD/MDD and hypertension/CAD are shown in Table 2.

**Table 2:**
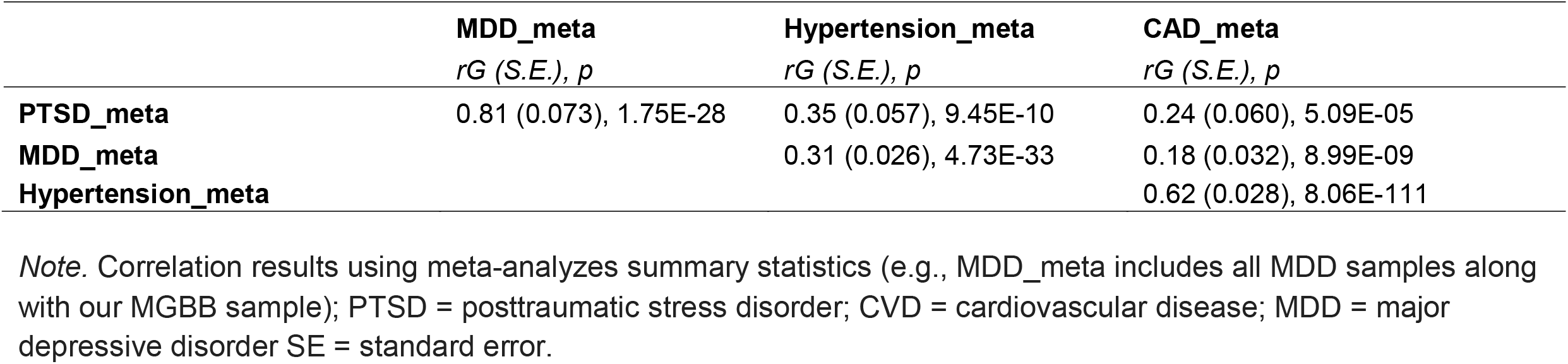
Genetic correlations among PTSD/MDD and CVD using meta-analyzed summary statistics.

### Improving genetic prediction of PTSD

Next, we sought to leverage these robust correlations to improve the genetic prediction performances for PTSD diagnosis. First, we combined only publicly available summary statistics using MTAG. The MGBB samples were not included in the base/training summary statistics, which were used as the target/testing dataset. As depicted in Table 3 and Figure 2, incorporating summary statistics for MDD and CVD resulted in improvement of both stratification as well as discrimination ability of the PRS, such that the prediction of PTSD diagnosis was significantly enhanced. Using only PGC-PTSD freeze-2 summary statistics resulted in AUC of 0.546 (CI: 0.532 - 0.560), while using summary statics that combines that combined PTSD, MDD, hypertension and CAD resulted in AUC of 0.592 (CI: 0.578 - 0.606). See Figure S2 for hypertension and CAD prediction performance.

**Table 3:**
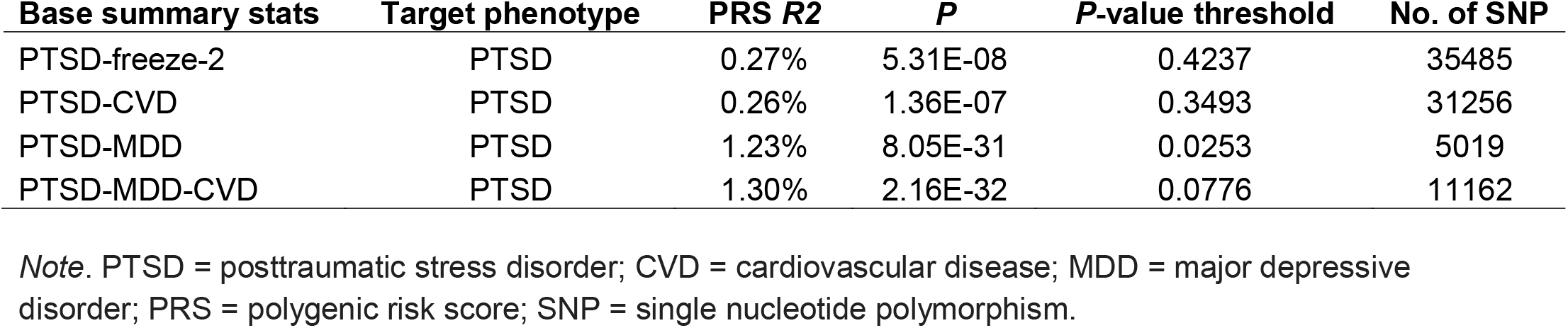
The performance of the PTSD polygenic risk score improves with incorporation of MDD and CVD summary statistics.

| **Base summary stats** | **Target phenotype** | **PRS $R^2$** | **$P$** | **$P$-value threshold** | **No. of SNP** |
| --- | --- | --- | --- | --- | --- |
| PTSD-freeze-2 | PTSD | 0.27% | 5.31E-08 | 0.4237 | 35485 |
| PTSD-CVD | PTSD | 0.26% | 1.36E-07 | 0.3493 | 31256 |
| PTSD-MDD | PTSD | 1.23% | 8.05E-31 | 0.0253 | 5019 |
| PTSD-MDD-CVD | PTSD | 1.30% | 2.16E-32 | 0.0776 | 11162 |

*Note.* PTSD = posttraumatic stress disorder; CVD = cardiovascular disease; MDD = major depressive disorder; PRS = polygenic risk score; SNP = single nucleotide polymorphism.

**Figure 2:**
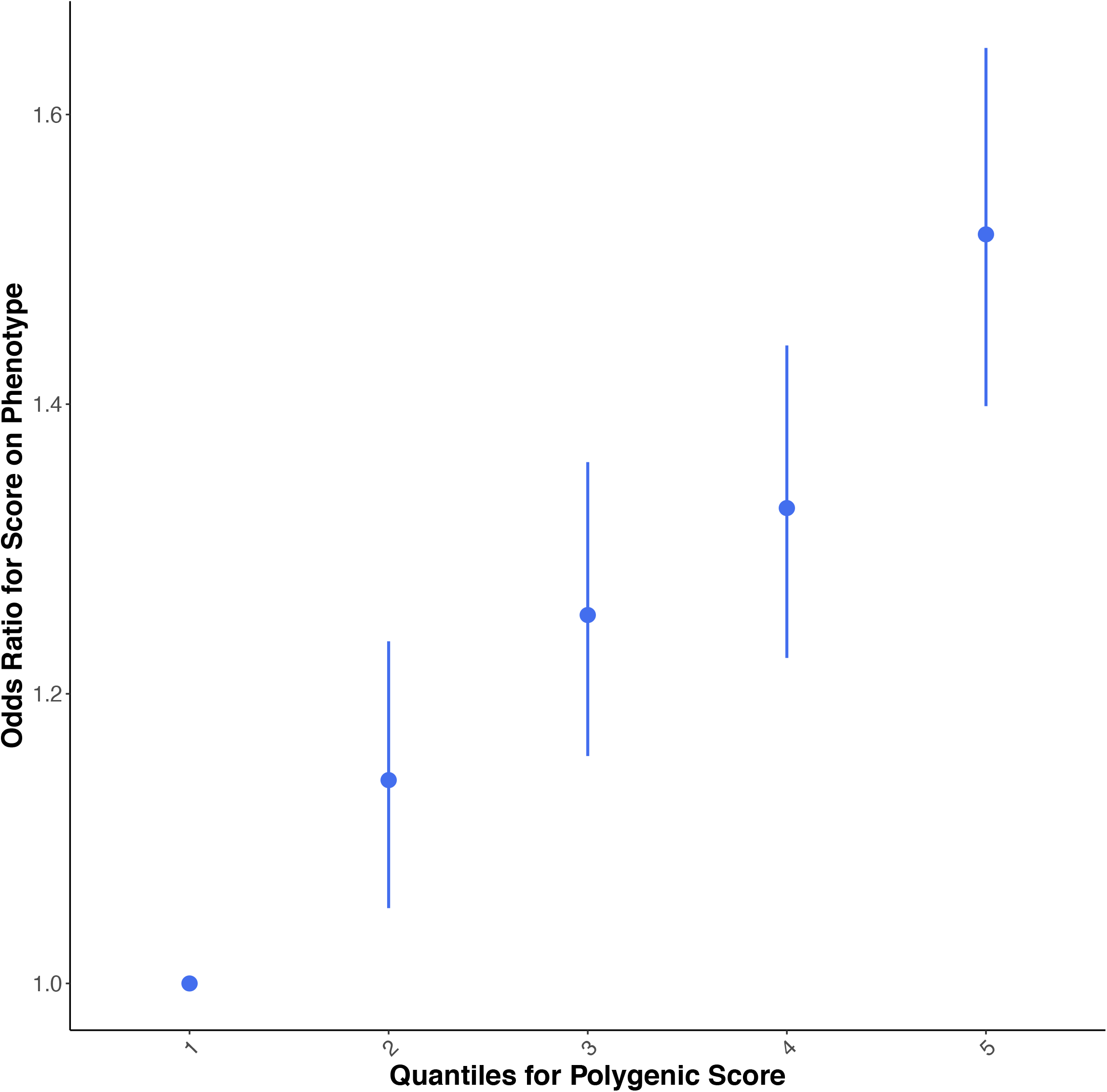
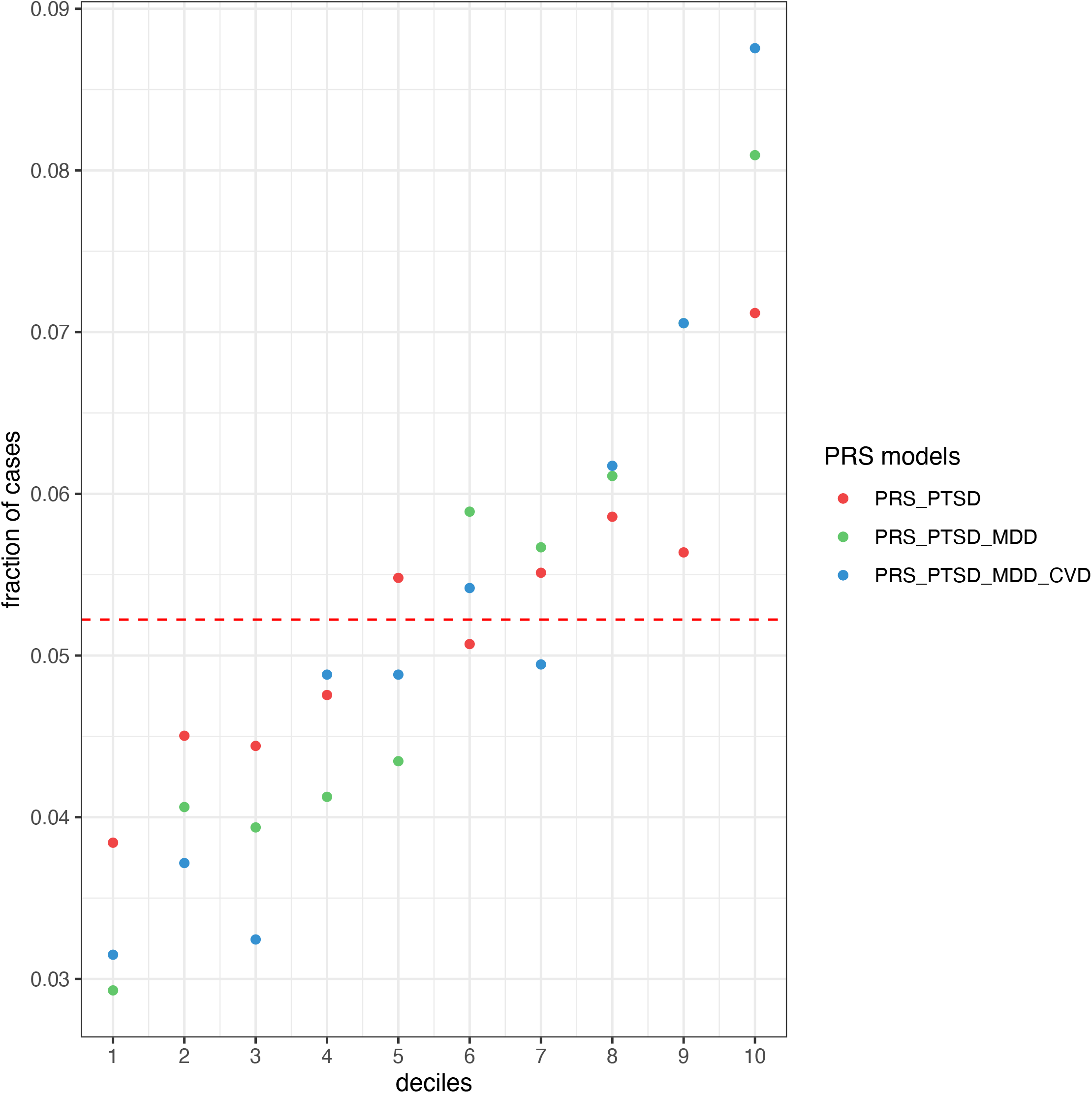
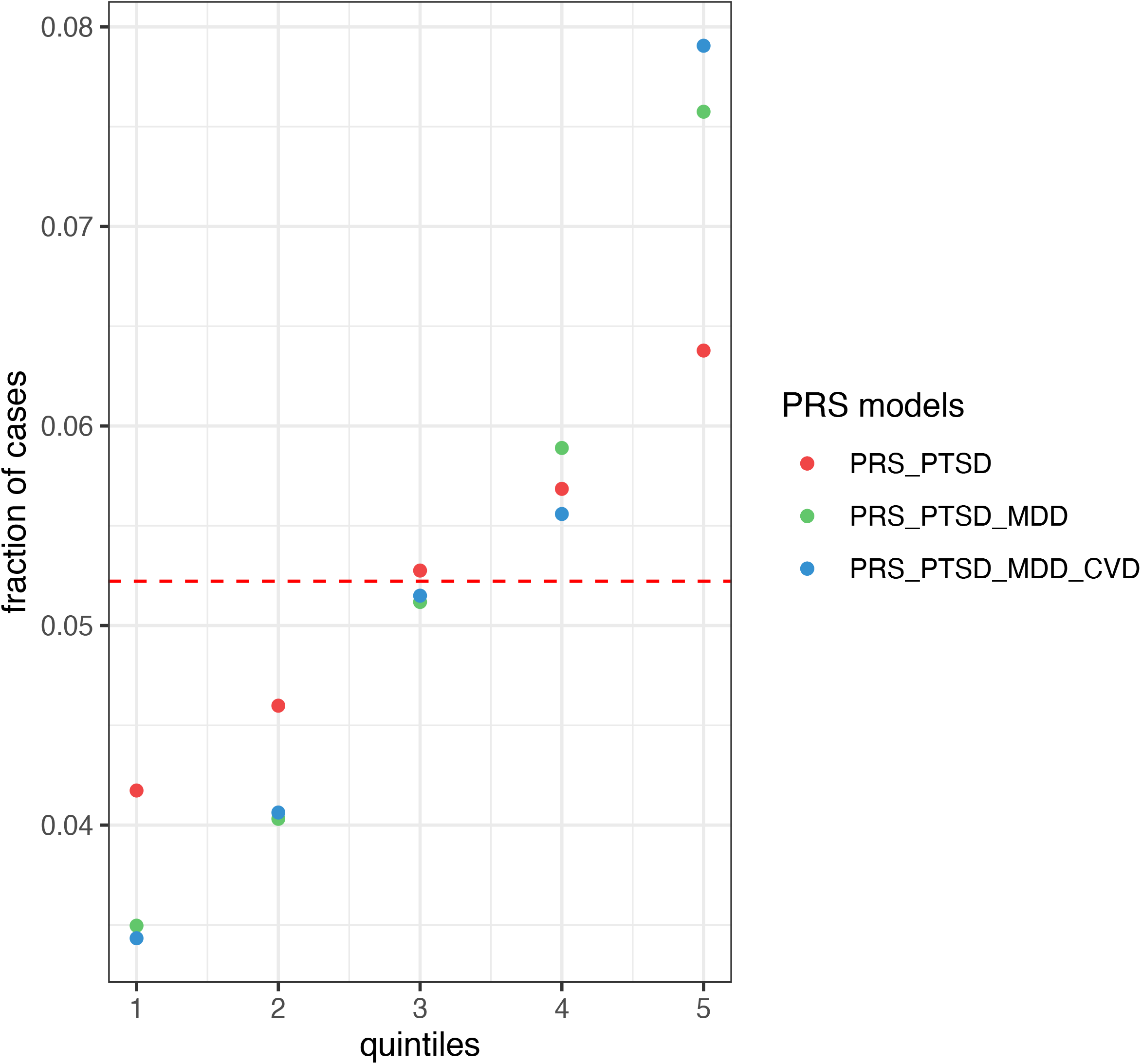
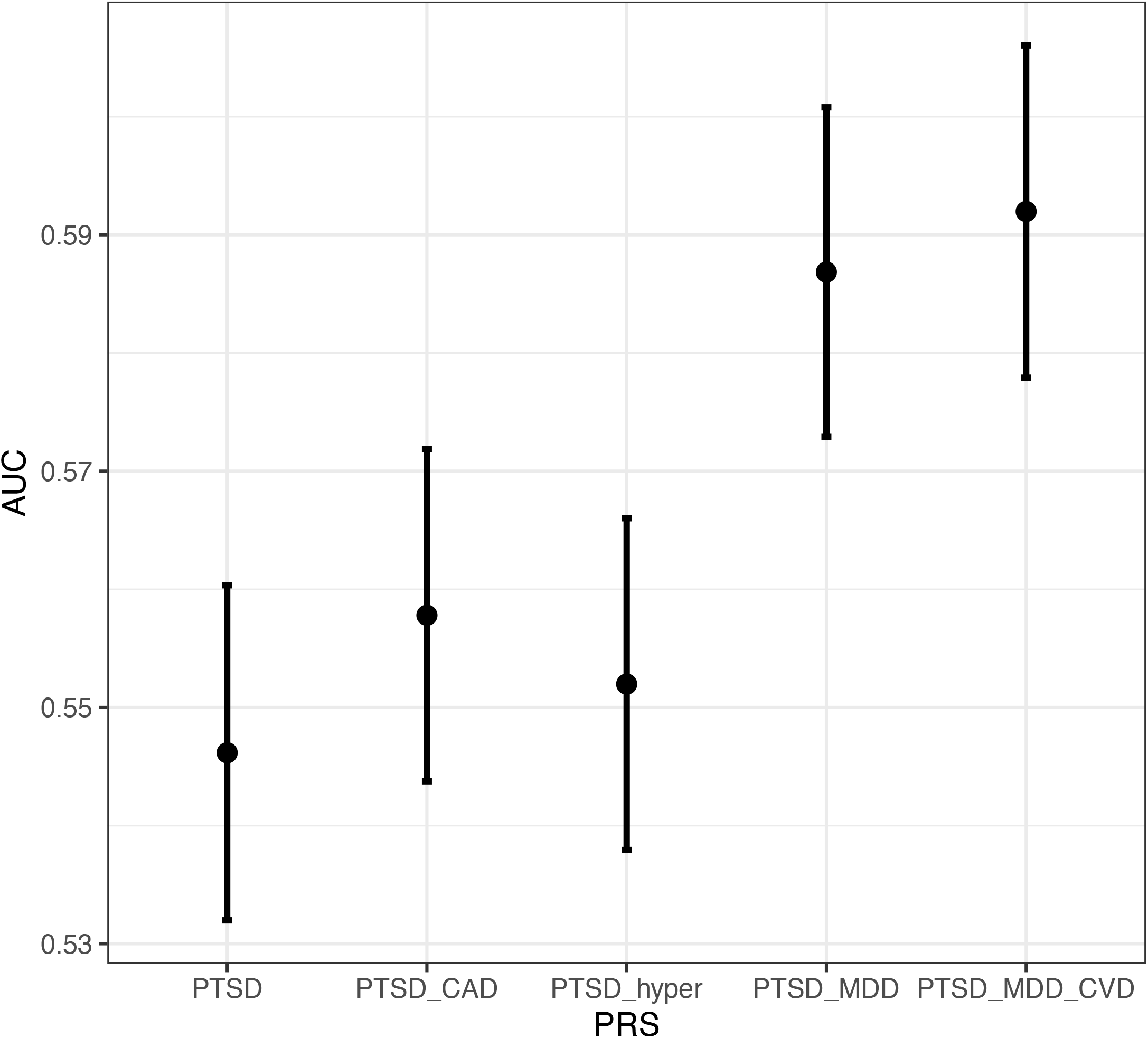
Stratification and discrimination ability of the polygenic risk score (PRS) constructed with various summary statistics. Figure 2a: Odds-ratio of PTSD diagnosis in PRS stratified risk groups. The first risk group is used as reference. Figure 2b: Fraction of participants diagnosed with PTSD in risk groups stratified into deciles (using PTSD, MDD, and CVD summary statistics). Figure 2c: Fraction of participants diagnosed with PTSD in risk groups stratified into quintiles (using PTSD, MDD, and CVD summary statistics). Figure 2d: Discrimination ability of the various PRS as measured by area under the curve (AUC), with greatest AUC for PTSD+MDD+CVD summary statistics.

### Mendelian randomization to determine causal PTSD-CVD link

Given that eight models were tested, we used a Bonferroni-corrected *p* value of 0.00625 (=0.05/8). All of the models with PTSD and MDD predicting hypertension and CAD were significant using this threshold: PTSD predicting hypertension (*β*=0.20, S.E.= 0.04, *p=5*.*97E-06*), PTSD predicting CAD (*β*=0.22, S.E.= 0.07, *p=9*.*22E-04*), MDD predicting hypertension (*β*=0.43, S.E.= 0.10, *p=5*.*97E-06*), and MDD predicting CAD (*β*=0.47, S.E.= 0.14, *p=9*.*22E-04*). Of the models in the reverse direction, only the model with CAD predicting PTSD was significant, but barely over the threshold of *p* < 0.00625 (*β*=0.31, S.E.= 0.11, *p=0*.*0041*). See Table S4 for a summary of all eight models. Overall, these findings provide greater support for a causal link in the direction from psychiatric disorders to hypertension and CAD, but not the reverse.

### Pathway enrichment analysis with shared genetic variants

A set of genetic variants at the significance threshold of *p* < 1.0E-5 (for MTAG-combined GWAS results with MGBB along with base summary statistics that combined PGC-PTSD, UKBB-depression, hypertension, and CAD) were used to perform pathway annotation and functional analysis with FUMA. This p-value threshold was used to include more genes and variants for pathway analysis, which is a common practice for these analyses^43^. In the first step, SNP2GENE, the set of variants are mapped to genes based on genetic coordinates. The 87 genomic risk loci included in the pathway analysis are shown in Figure S3. The 8,734 SNP’s falling in these genomic risk loci are mapped to 198 genes. In the second step, GENE2FUNC, the prioritized genes are used as input to the second step to perform functional annotation. Top significantly enriched gene ontology biological processes in the set of genes included those involved in the regulation of postsynaptic structure (*p= 0*.*00001, adjP=0*.*005*), synapse organization (*p= 0*.*0001, adjP=0*.*02*), and interleukin-7 (IL-7) mediated signaling pathways (*p= 0*.*0002, adjP=0*.*03*). In addition to the pathways, tissue specificity was assessed using differentially expressed genes (comparing a given tissue label versus all other labels). However, no statistically significant result was found (see Figure S4).

## Discussion

This is the first study to examine the shared genetic susceptibility of PTSD and CVD utilizing large-scale genome-wide cross-trait analysis from both PTSD and CVD GWAS summary statistics, which were derived from a total of 1.3 million samples, and 31,758 genotyped samples that were used as target/test dataset. We identified significant genetic correlations between PTSD and CVD, as well as MDD, and Mendelian randomization analyses provided support for a causal link from PTSD to hypertension and CAD, but not the reverse. We were also able to significantly improve the genetic prediction of PTSD by incorporating summary statistics from CVD and MDD. Further, pathway enrichment analyses indicated that genetic variants involved in shared PTSD-CVD risk included those involved in postsynaptic structure, synapse organization, and immune function. These findings suggest that PTSD and CVD risk identification may be improved by their mutual consideration, which can be used to improve PTSD treatment selection.

A preponderance of epidemiological data has suggested that individuals with PTSD are more likely to experience CVD.^2,3^ As identified by the NHLBI working group^6^ on PTSD-CVD risk, large-scale studies are needed to better understand the shared genetic risk of these diseases. Here we provide the first evidence of shared genetic risk for PTSD and CVD using summary statistics from both literatures. We also demonstrated significant genetic associations of PTSD and CVD with MDD, which is consistent with prior research supporting comorbidity of MDD with both PTSD and CVD.^29^ While the overlap of mental disorders is increasingly accepted in psychiatry,^44^ our results suggest that two highly impairing psychiatric conditions also overlap with a set of cardiovascular diagnoses thought to have more discrete origins. This provides further evidence that psychiatric and physical health conditions are not only common in phenotypic presentation, but that they are genetically linked.

Our Mendelian randomization analyses found evidence that genetic risk for PTSD and MDD exerted a causal effect on hypertension and CAD risk. One of the primary mechanisms implicated in the link between PTSD/MDD and CVD is elevated sympathetic arousal that leads to hypertension, both directly (i.e., chronically elevated BP due to stress) and via the renin-angiotensin system (i.e., elevated BP due to renin and angiotensin-II release that causes vasoconstriction). Our findings provide support for this mechanism, however, we cannot exclude the possibility that confounding factors or mediating effects (e.g., diet, smoking) are responsible for these associations, which should be examined in future research. Our pathway enrichment analyses suggested that genetic variants involved in postsynaptic structure, synapse organization, and immune function (IL-7) may be relevant to the shared genetic risk of PTSD and CVD. While IL-7 has not previously been studied in the context of PTSD, other immune markers and genes (e.g., IL-6, IL-8) have been implicated in PTSD and it has been suggested that impaired immune function may be a mechanism underlying the PTSD-CVD link.^5,26^ Given that IL-7 has been implicated in hypertension and atherosclerosis^45,46^, this further implicates the role of immune function in the shared genetic risk of PTSD and CVD, and it suggests that additional study of these genetic variants is warranted.

The primary clinical implication of this work is improved risk identification, which can be used to enhance PTSD treatment and decrease CVD risk. For example, identifying individuals with shared PTSD and CVD risk (and knowing that PTSD is a specific risk factor for CVD) will allow clinicians to select PTSD interventions that are known to improve aspects of cardiovascular function. There is preliminary evidence that forms of cognitive-behavioral therapy (the first line treatment for PTSD) improve cardiovascular function,^46^ but this research is still in its early stages. On the other hand, psychiatric and CVD medications have well-documented effects on cardiovascular function,^48,49^ but few studies have tested the effects of these medications on subsequent CVD risk in PTSD populations. Medications targeting the renin-angiotensin system (e.g., ACE-inhibitors, beta-blockers) have demonstrated efficacy in rodent models, but research in humans with PTSD has been mixed.^50, 51^ A next step in this line of work is to determine if existing cognitive-behavioral and pharmacological treatments actually reduce CVD risk in PTSD, and to determine if they are more efficacious for individuals with high genetic risk for both PTSD and CVD. An additional and more immediate next step is to include PTSD as an indicator of CVD risk in routine care (e.g., during hypertension screening).

While our use of a large biobank sample is a strength of the current study, it also has inherent limitations. For example, PTSD diagnosis was based on single ICD-10 codes, which were not necessarily the result of standardized assessments (e.g., the Clinician Administered PTSD Scale^52^). This approach, while necessary for large-scale data analyses, is not as reliable as using clinician-administered assessments of PTSD as we cannot determine whether some PTSD diagnoses were given incorrectly. Another limitation is that the MGBB sample is comprised primarily of White, non-Hispanic individuals. As a result, over 87% of participants were of EUR ancestry and analyses were conducted only on these individuals. While this approach minimized the effect of population stratification, it means that our findings may not replicate to other racial/ethnic/ancestry groups. Finally, it is important to note that the relationship between PTSD and CVD is likely mediated, at least in part, by behavioral risk factors, such as diet, exercise, smoking, and alcohol/substance use. These risk factors are independently associated with CVD risk and cardiometabolic diseases that are linked with CVD (e.g., Type-2 diabetes mellitus). Thus, a limitation to the current study is the lack of data on these behavioral variables that likely influence the pathways through which PTSD may lead to CVD. Future research using multivariate Mendelian randomization approaches are needed to further assess the role of possible confounding or mediating factors in the PTSD-CVD link. Similarly, it will be critical for future studies to test whether the PTSD and MDD correlations with CAD depend on hypertension, and whether PTSD and MDD each have unique risk associated with hypertension and CAD independent of one another.

The link between PTSD and CVD has been well-established from an epidemiological standpoint. As identified by the 2018 NHLBI working group,^6^ large-scale genetic studies are needed to improve characterization of PTSD and CVD risk, which will subsequently improve risk management and treatment. We directly addressed some of these needs by utilizing a large biobank sample (*N* = 36,412) and by leveraging large-scale GWAS summary statistics from both the PTSD and CVD literatures with 1.3 million samples. Our results indicate that 1) there is substantial genetic overlap between PTSD and CVD, 2) PTSD and MDD may be risk factors leading to the development of hypertension and CAD, and 3) the genetic prediction of PTSD risk is improved by the consideration of polygenic risk for CVD and MDD. Future studies of genetic risk and diagnostic prediction would benefit from incorporating this polygenic risk approach.

## Data Availability

All data produced in the present work are contained in the manuscript.

## Figure titles

Supplementary Figure 1: Thresholding step with MTAG summary statistics for PTSD polygenic risk score (PRS).

Supplementary Figure 2: Improvement in the prediction of coronary artery disease (CAD) and hypertension using PTSD and MDD summary statistics.

Supplementary Figure 3: Genomic risk loci included in pathway analysis.

Supplementary Figure 4: Tissue specificity of pathway analysis.

Supplementary Figure 5: MGBB_CAD_manhattan

Supplementary Figure 6: MGBB_ CAD_qq

Supplementary Figure 7: MGBB_ CHF_manhattan

Supplementary Figure 8: MGBB_ CHF_qq

Supplementary Figure 9: MGBB_ DiastolicAvg_manhattan

Supplementary Figure 10: MGBB_ DiastolicAvg _qq

Supplementary Figure 11: MGBB_ Hypertension_manhattan

Supplementary Figure 12: MGBB_ Hypertension_qq

Supplementary Figure 13: MGBB_ MDD_manhattan

Supplementary Figure 14: MGBB_ MDD_qq

Supplementary Figure 15: MGBB_ MI_manhattan

Supplementary Figure 16: MGBB_ MI_qq

Supplementary Figure 17: MGBB_ PTSD_manhattan

Supplementary Figure 18: MGBB_ PTSD_qq

Supplementary Figure 19: MGBB_ PulseAvg_manhattan

Supplementary Figure 20: MGBB_ PulseAvg_qq

Supplementary Figure 21: MGBB_ SystolicAvg_manhattan

Supplementary Figure 22: MGBB_ SystolicAvg_qq

## Supplementary Material

### Phenotype data

Diagnostic data was extracted from the MGBB and ICD-10 codes and curated disease populations were used to determine diagnosis.^31,32^ Curated Disease Populations are collected by a validated phenotype algorithm with a positive predictive value of 0.90. These algorithms rely on coded diagnosis as well as natural language processing terms extracted from clinical narratives. Predictive features for each characterization were identified using an automated feature extraction protocol, which identifies comorbidities, symptoms, and medications. Concepts were screened based on frequency in patient clinical notes. Billing diagnosis and prescriptions were also included in algorithms.^31,32^ Patients with lifetime history of congestive heart failure, coronary artery disease, hypertension, ischemic stroke, and depression as determined by Curated Disease Population status were considered positive for these traits in our dataset. Curated Disease Populations were not available for PTSD or myocardial infarction.

### Curated Disease Population Algorithms

Model definitions with features and betas: Below are the feature weights of the final hypertension phenotype algorithms. The weights below are used to derive a predicted probability of hypertension or no hypertension for every Biobank participant.

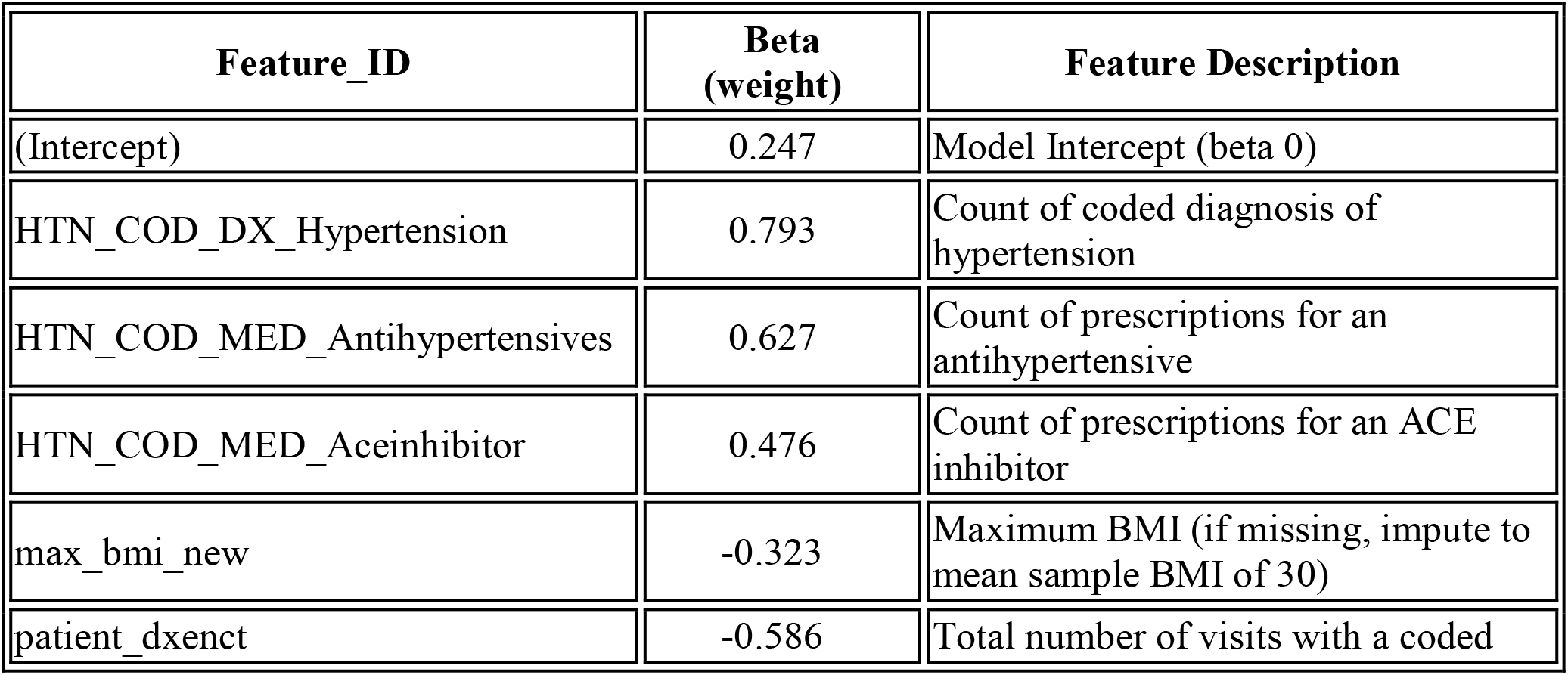
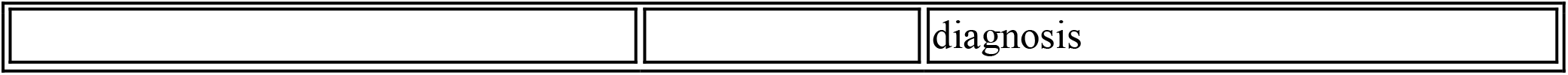

Below are the feature weights of the final CAD phenotype algorithms. The weights below can be used to derive a predicted probability of CAD or no CAD for any Biobank participant.

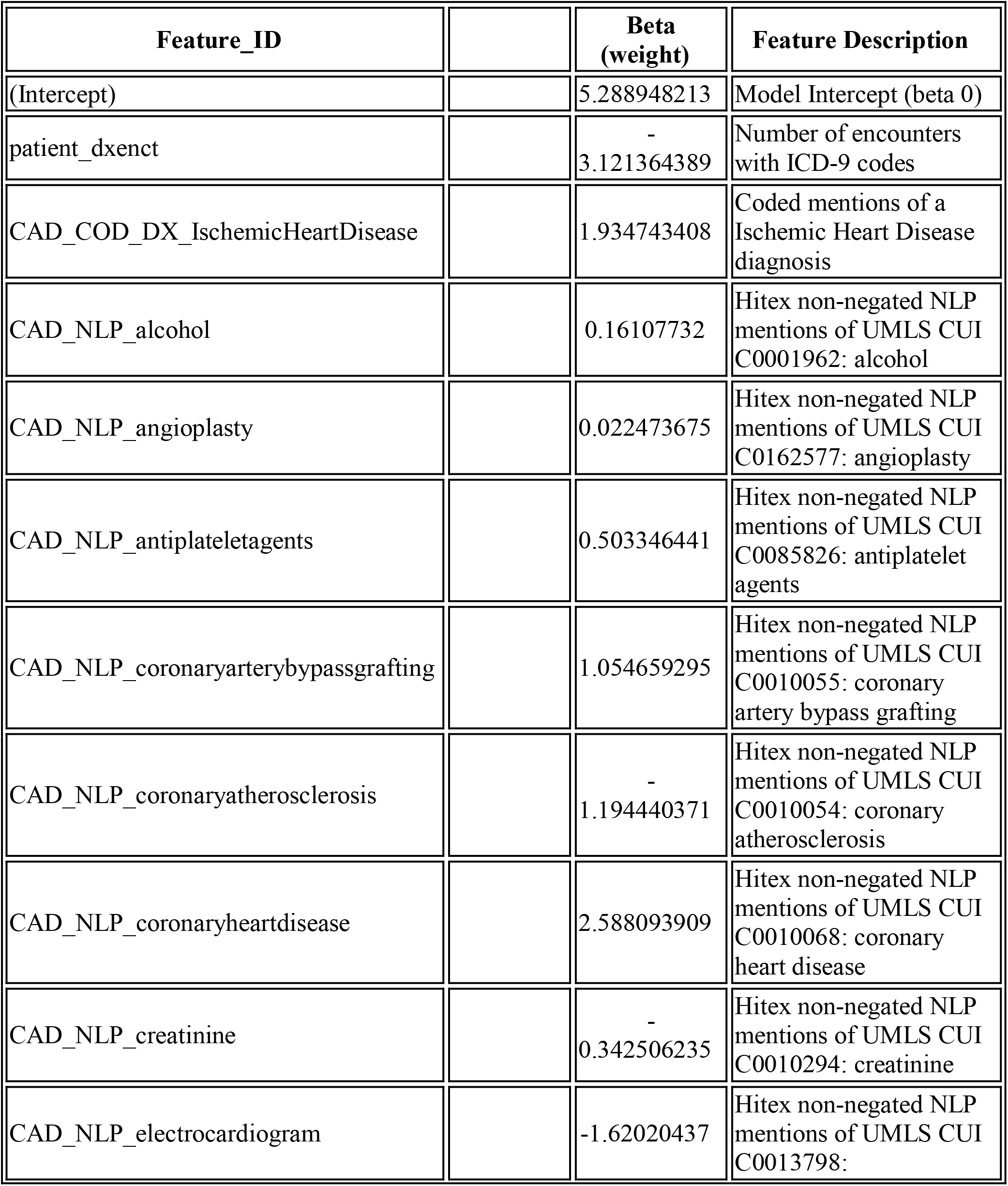
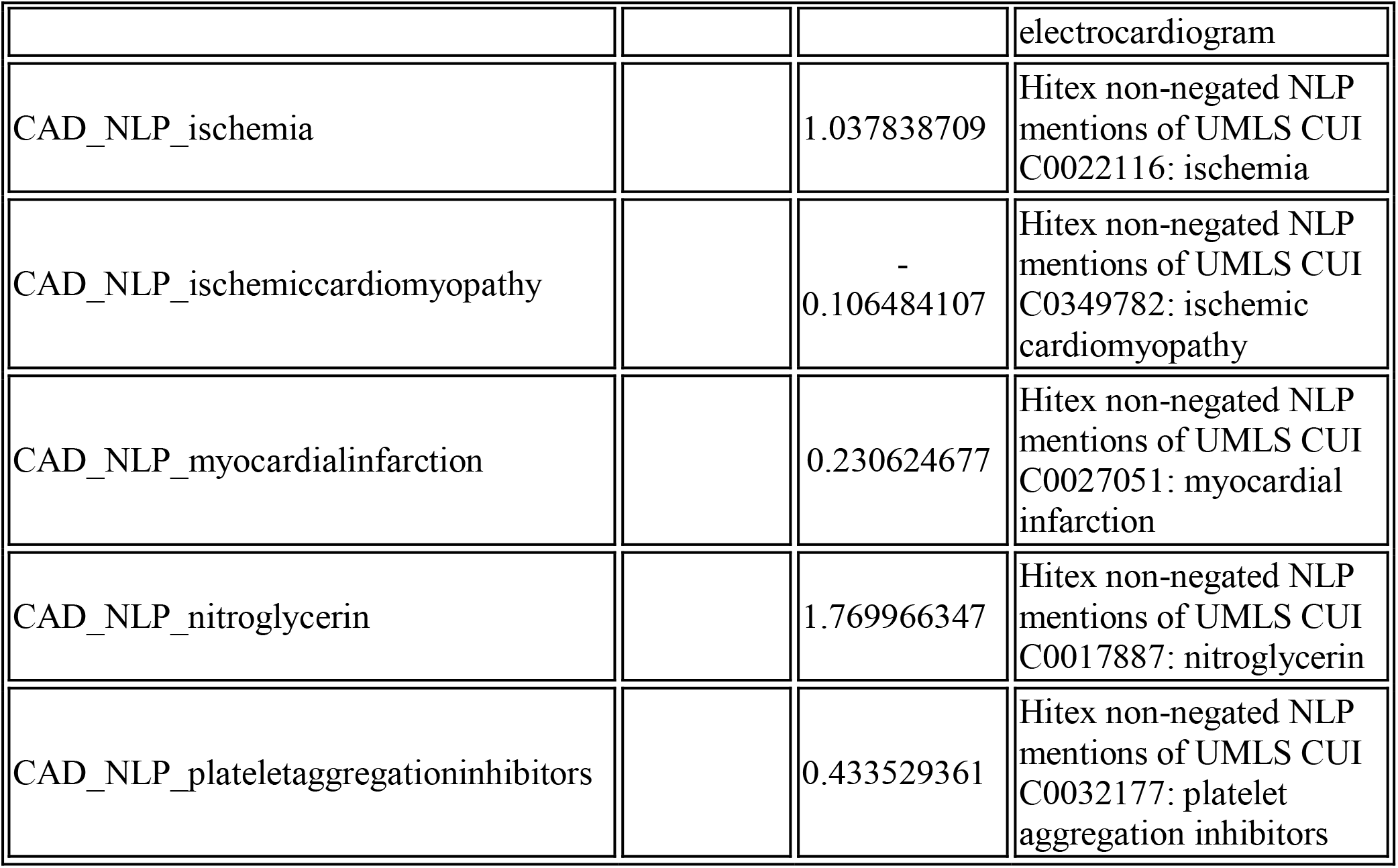

Below are the feature weights of the final depression phenotype algorithms. The weights below are used to derive a predicted probability of depression or no depression for every Biobank participant.

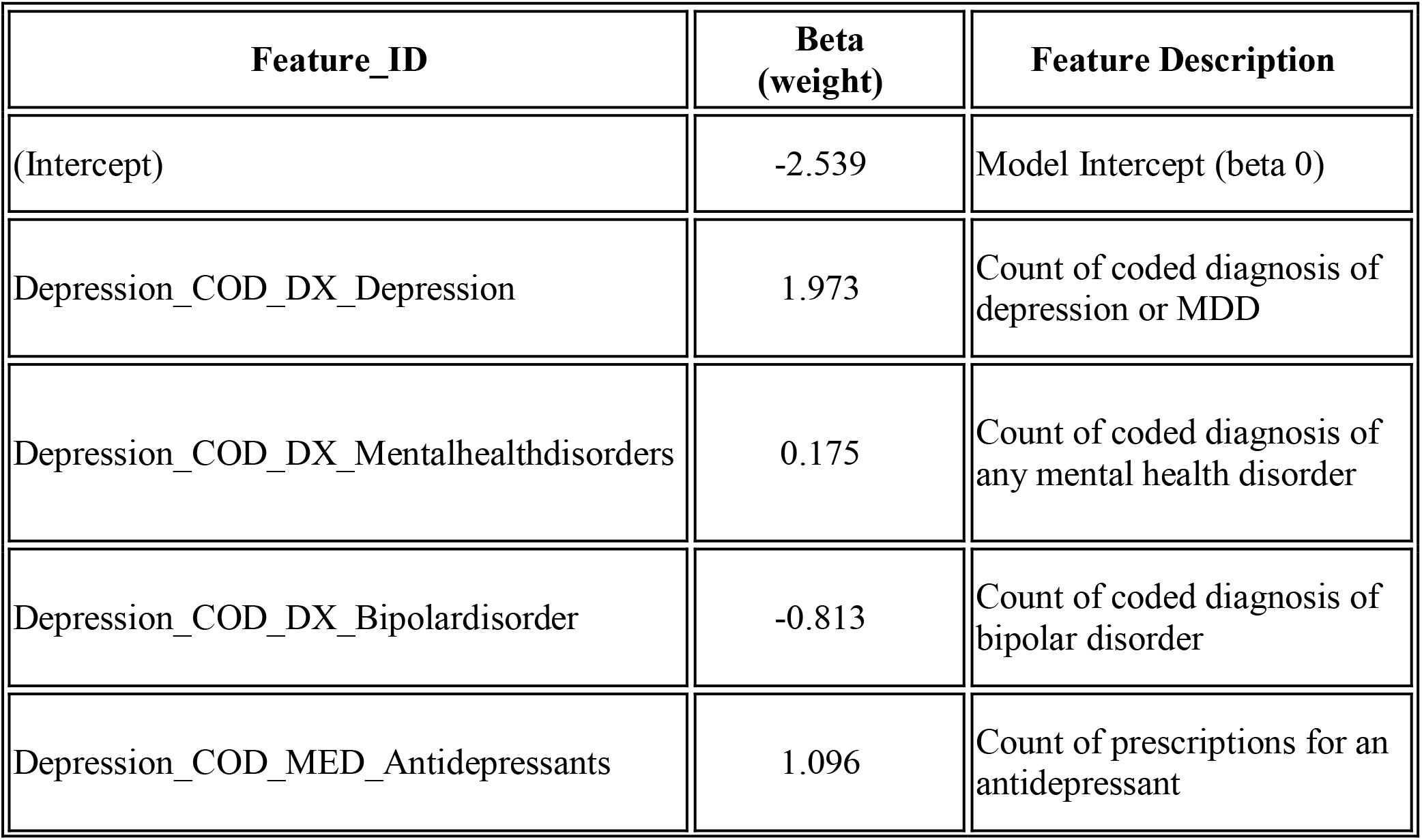
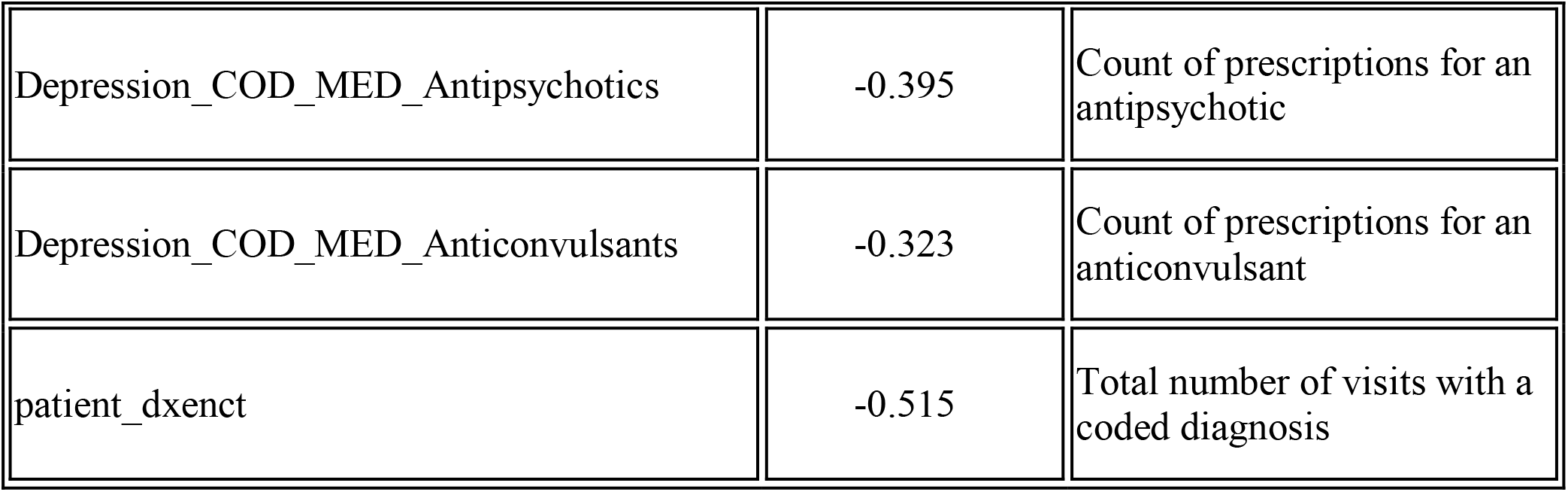

Below are the feature weights of the final CHF phenotype algorithms. The weights below can be used to derive a predicted probability of CHF or no CHF for any Biobank participant.

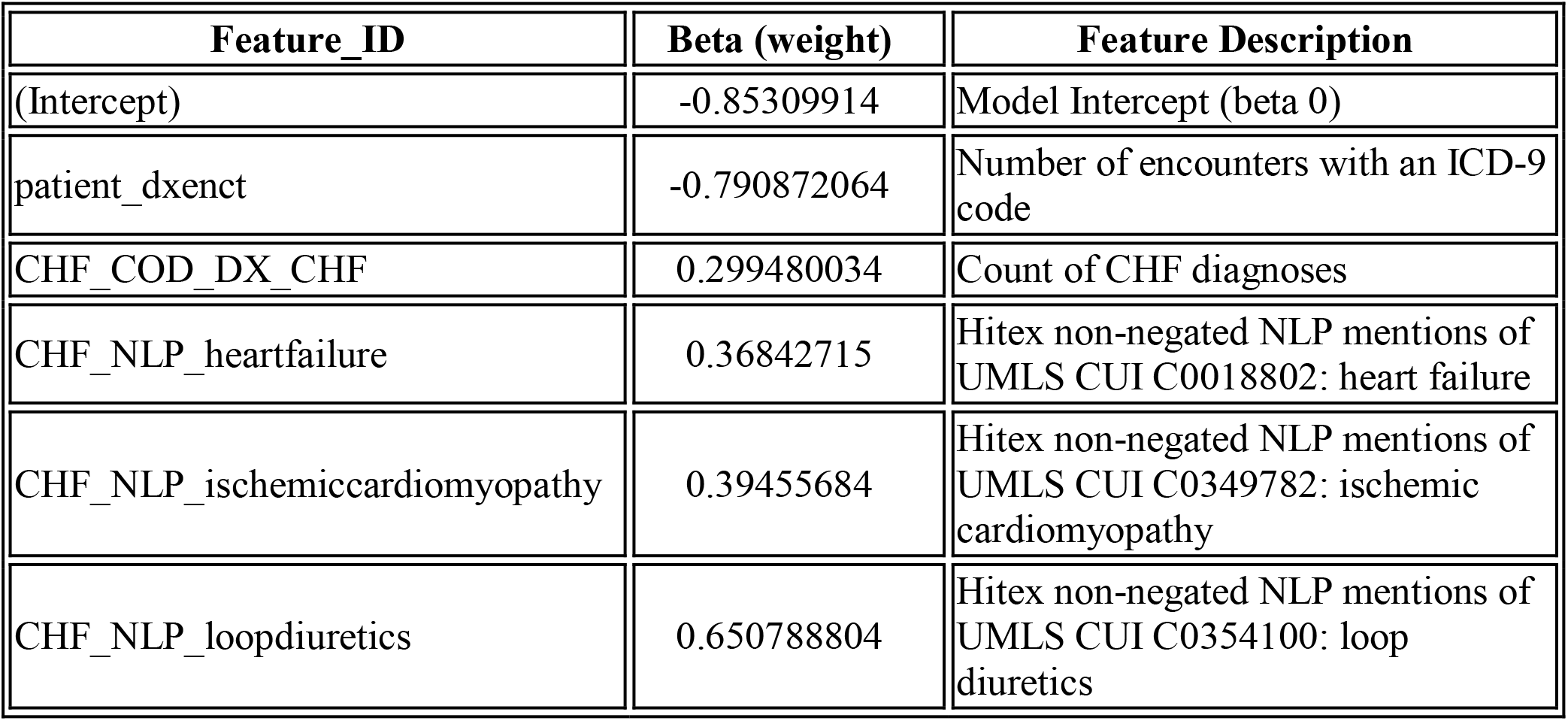

Below are the feature weights of the final Ischemic stroke phenotype algorithms. The weights below are used to derive a predicted probability of ISTR for every Biobank participant.

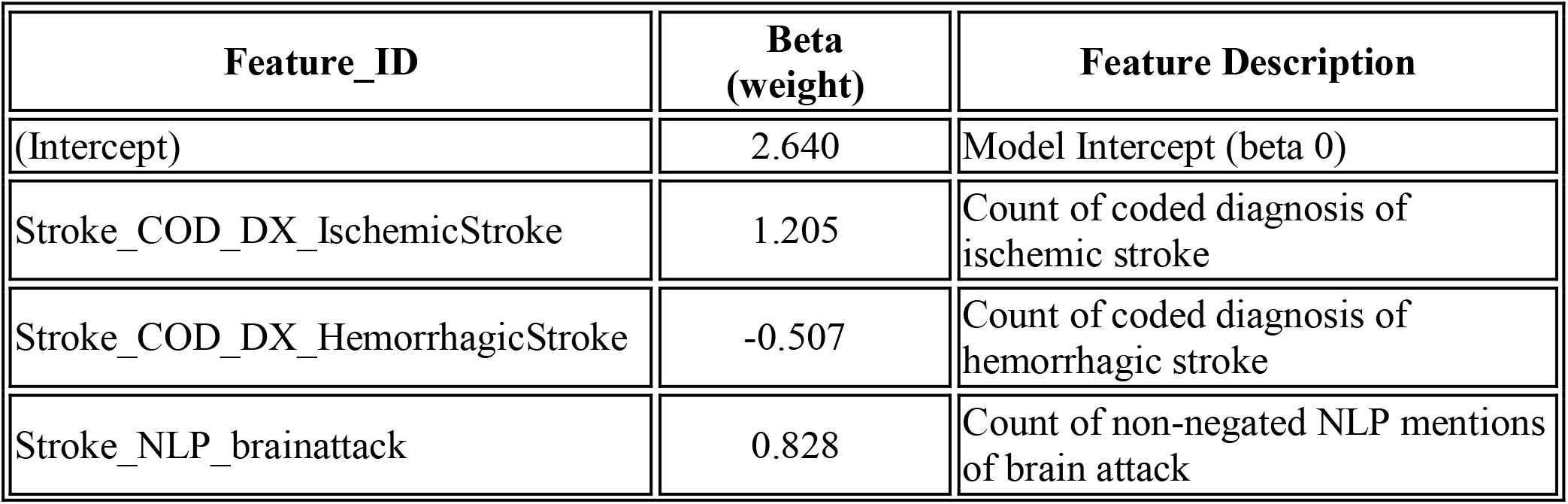
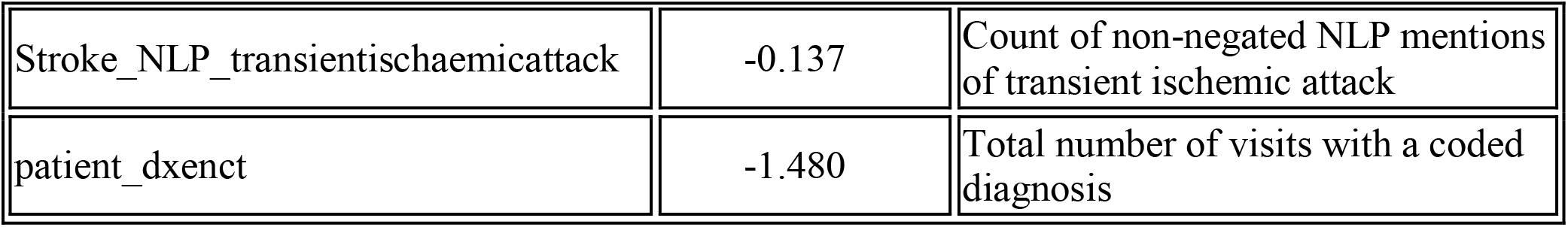

### Genetic data

De-identified genetic data was requested from the MGBB. Whole blood samples are collected from participants during research draws or at the time of clinical draws. Samples are genotyped in batches using three versions of Illumina SNP array. Imputed genotype data from Multi-Ethnic Genotyping Array (MEGA), Expanded Multi-Ethnic Genotyping Array (MEGA Ex), and Multi-Ethnic Global (MEG) BeadChip were included in this analysis. Imputation was performed using the Michigan Imputation Server with a Minimac3. Only variants on the 22 autosomal chromosomes were considered. Data were obtained in 8 batches/sub-cohorts: Batch 1: MEG_A1_A (*n*=4780), Batch 2: MEG_A1_B (*n*=5020), Batch 3: MEG_C (*n*=5492), Batch 4: MEG_D (*n*=5146), Batch 5: MEG_E (*n*=4850), Batch 6: MEG_X1 (*n*=866), Batch 7: MEGA (*n*=4924), Batch 8: MEGAEX (*n*=5344).

### Preprocessing and QC of genetic data

QC was performed on each batch individually and merged together, resulting in a total of 36,422 individuals and 9,036,179 variants after imputation. Unique ID’s (rsID’s) of variants were inserted based on chromosomal locations from the dbSNP database. Variants were filtered with two more criteria: minor allele frequency (MAF>0.01) and Hardy-Weinberg equilibrium (HWE p-value>1E-6). This resulted in 6,001,335 SNPs.

### Mendelian randomization analyses

Genetic correlations and phenotypic comorbidity between PTSD/MDD and hypertension (and cardiovascular illnesses in general) have been repeatedly reported. In the current manuscript we demonstrated significant genetic correlations between the two psychiatric disorders and cardiovascular illnesses with a robust statistical evidence. These correlations observed at various levels may stem from one of the following four scenarios/mechanisms.

- <u>Scenario 1</u>: Onset of MDD/PTSD causes higher predisposition to hypertension and other cardiovascular illnesses. This could possibly be mediated by lifestyle changes (such as smoking and less exercise/mobility) and subsequent decline in anthropometric measurements (such as higher BMI).
- <u>Scenario 2</u>: Existence of cardiovascular illnesses leads to elevated predisposition to MDD/PTSD.
- <u>Scenario 3</u>: Both MDD/PTSD and cardiovascular illness are caused by shared genetic variants. This shared genetic underpinning can possibly influence distinct pathways/processed in the two groups of disorders. This is commonly known as horizontal pleiotropy.
- <u>Scenario 4</u>: MDD/PTSD and hypertension are caused by separate genetic variants that happen to be in linkage disequilibrium with each other.

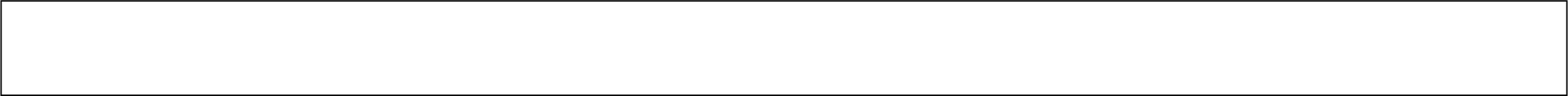

It is important to note previous publications suggesting positive causal effect of MDD on cardiovascular diseases (Scenario 1 above) [PMID: 33032663], [PMID: 33372528]. Another interesting observation is the significant difference in age distribution between the two group of disorders. In the MGBB data, the median age for PTSD-positive and MDD-positive patients are 51 and 62; while the median age for hypertension and CAD positive patients are 70 and 75, respectively. Hence, those diagnosed with cardiovascular illnesses are, on average, significantly older than those diagnosed with the psychiatric disorders. Even if this is not observed on a longitudinal follow-up observation, it is suggestive of the sequence of which disorder is likely to be diagnosed first.

Here, we hypothesize that there is a causal link from MDD/PTSD to hypertension. The gold standard for inferring a causal link is Randomized control trial (RCT). However, in most situations, RCT is not feasible and requires a long timeframe to implement. This holds true in our current dataset as well. A feasible alternative that is cost-effective and implementable using our current study data is Mendelian Randomization (MR) [PMID: 34698778]. MR can be thought of as a ‘randomized trial’ where randomization is done at birth, where predisposing genetic variants are assorted randomly from parents to offspring [PMID: 19509388]. In other words, the predisposing genetic variants can be thought of as proxies for randomized intervention (assuming random mating).

MR is a special case of what is commonly known as instrumental variable analysis where genetic variation is used as the instrumental variable [PMID: 26282889]. For a variable to be used as an instrumental variable (genetic variants in MR case), three main assumptions should hold true: (1) Relevance: the genetic variant is associated with the exposure, (2) Exclusion restriction: the genetic variant does not affect the outcome directly, and (3) Random assignment: the genetic variant does not affect the outcome through confounding variables. The main goal is to infer a causal link from the exposure variable to the outcome variable.

To test the causal link from MDD/PTSD to hypertension using MR, MDD/PTSD is our exposure variable and hypertension is the outcome variable. There are two possible choices for genetic variants as instrumental variables. The first and more common approach is to select few individual variants that are significantly associated with the exposure. MR analysis is conducted on each genetic variant as the instrumental variable and the results are combined afterwards. The second approach is to use polygenic risk scores as instrumental variables. The use of polygenic scores instead of using multiple individual variants (by far the most common approach) has been shown to mitigate instrumental variable bias (PMID:24062299).

Here, we use polygenic scores computed in the current manuscript to conduct the MR procedure. We implemented a procedure commonly known as two stage least square regression (2SLS) [PMID: 24114802]. In the first stage, the exposure (binary MDD label) is regressed against MDD polygenic risk score and along with confounders we want to control for (age and the first five principal components). Using this regression model, predicted value of the exposure is computed (MDD_hat). In the second stage, the outcome variable (hypertension binary label) is regressed against the predicted exposure variable (MDD_hat) and the potential confounding variables (age and the first five principal components). Then, (epidemiological) causality is inferred based on the significance of the coefficient of the predicted exposure variable (MDD_hat). The result of the statistics of the coefficient is shown below.

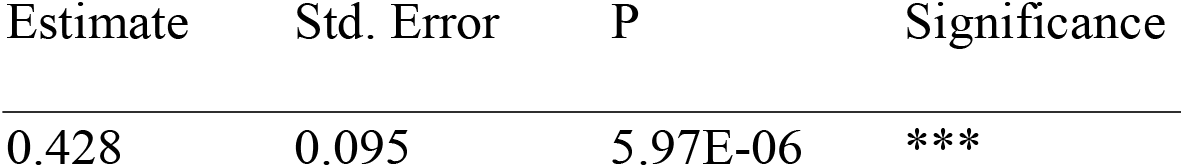

Therefore, we can infer that MDD onset is linked to hypertension diagnosis. We implemented a similar MR procedure for three other models. The results are shown in the table below.

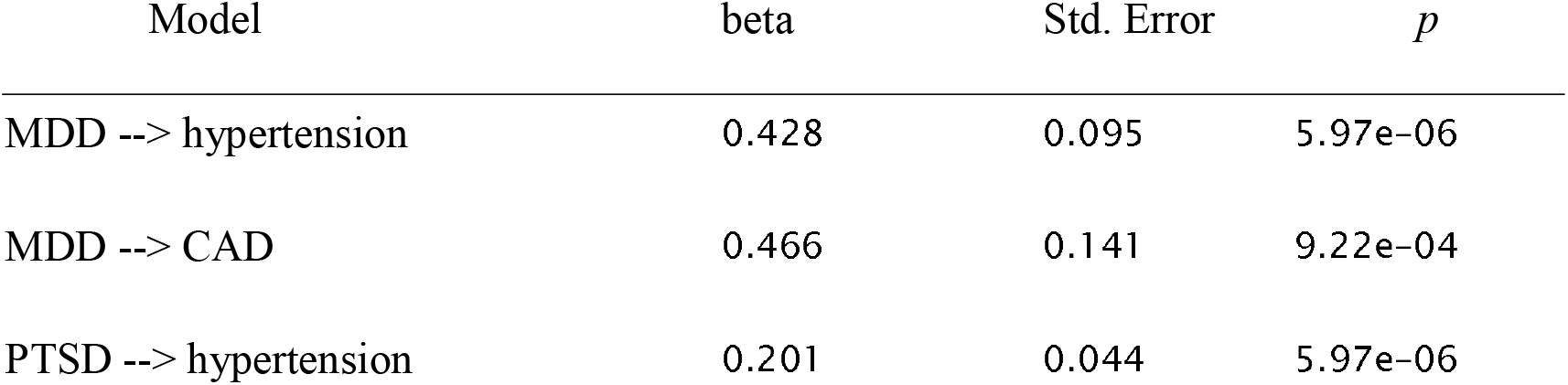
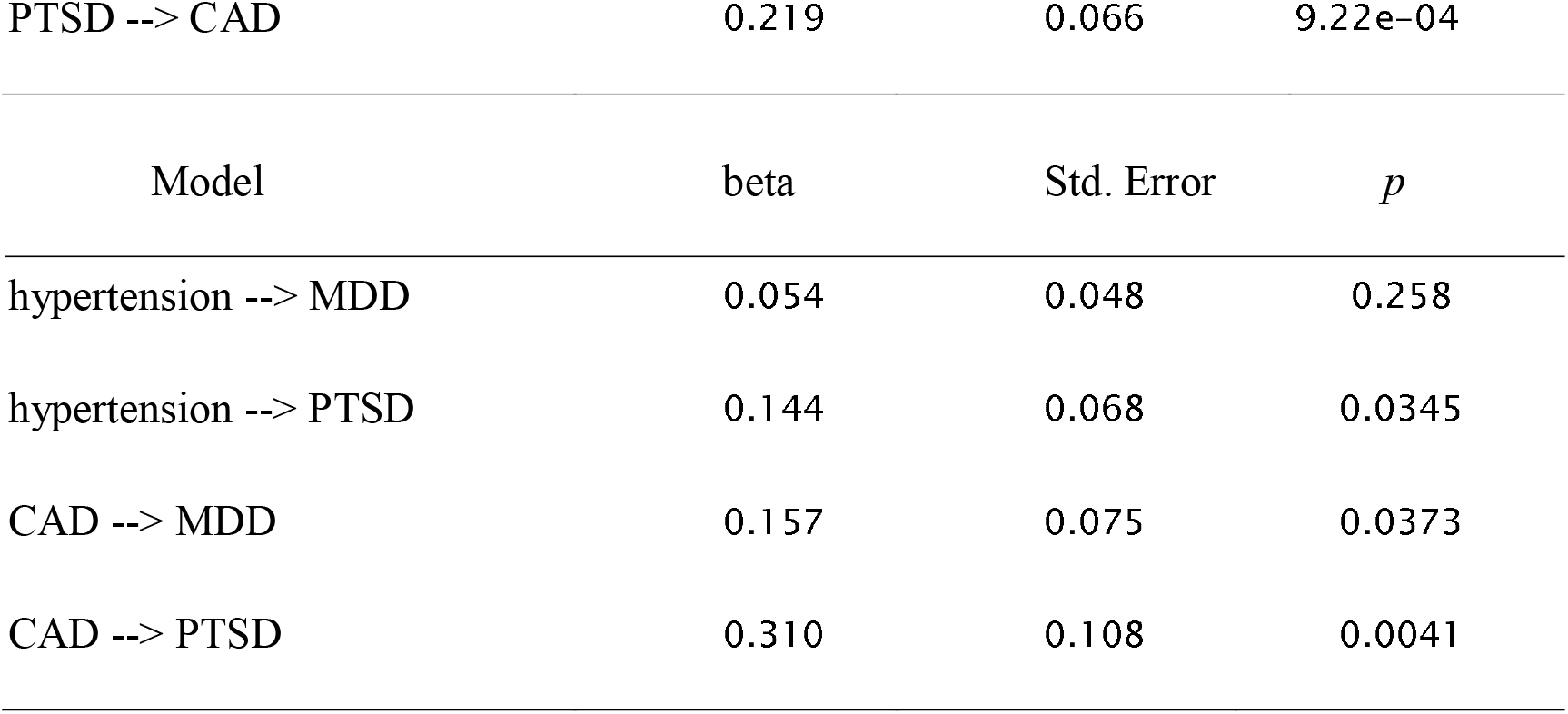

As shown, our analysis provides support for a causal link from psychiatric disorders to cardiovascular illnesses, but not the other way around.

Note that all analysis is only for EUR sub-population of the MGBB dataset. In the future, similar analysis on sub-populations (based on ancestry, gender and age) needs to be conducted. All analysis is done on R statistical software.

### CRP analyses

We investigated genetic level correlations between the two groups of phenotypes and the largest publicly available CRP GWAS summary statistics (PMID: 33462484). As expected, CRP level in the blood is genetically correlated with both group of phenotypes. The main result is summarized below.

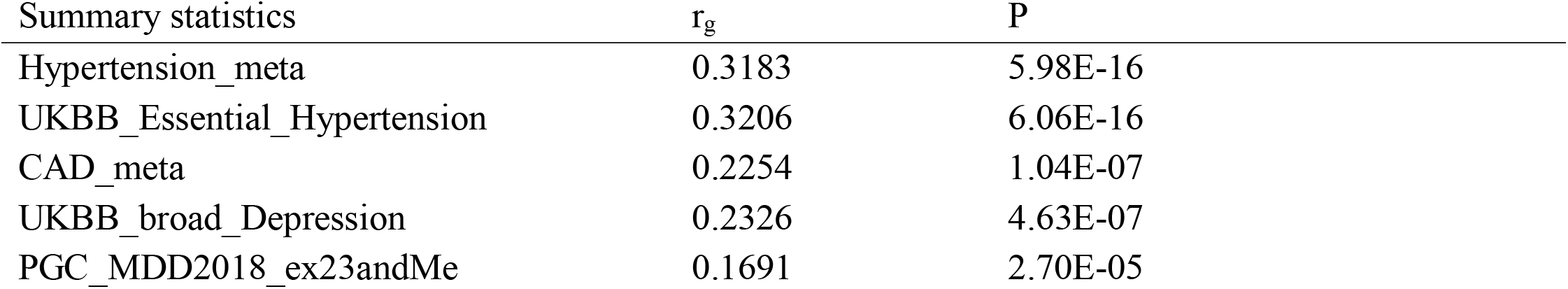
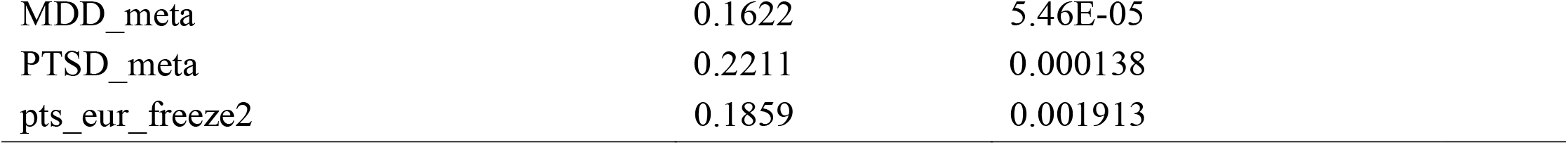

## Supplementary Tables and Figures

**Supplementary Table 1:**
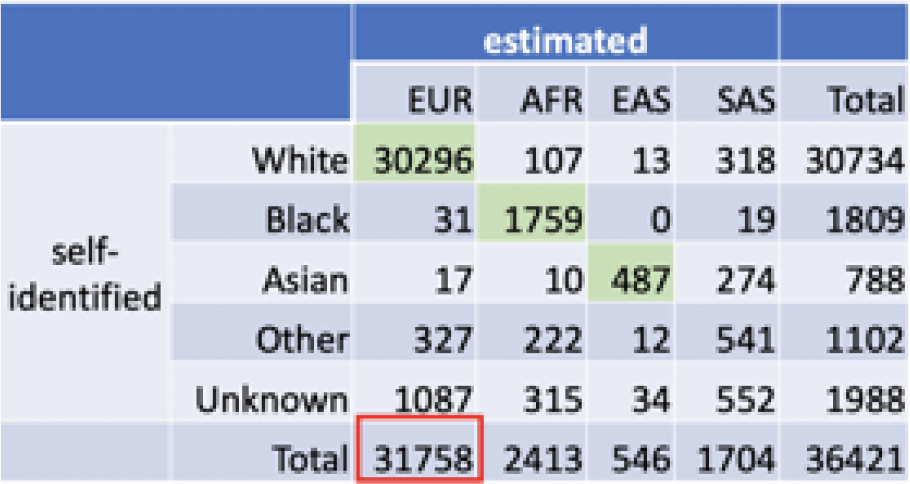
Self-identified and genetically predicted ancestry composition. Genetic analysis was done with those participants predicted to have EUR ancestry.

|  |  | estimated |  |  |  |  |
| --- | --- | --- | --- | --- | --- | --- |
|  |  | EUR | AFR | EAS | SAS | Total |
| self-identified | White | 30296 | 107 | 13 | 318 | 30734 |
|  | Black | 31 | 1759 | 0 | 19 | 1809 |
|  | Asian | 17 | 10 | 487 | 274 | 788 |
|  | Other | 327 | 222 | 12 | 541 | 1102 |
|  | Unknown | 1087 | 315 | 34 | 552 | 1988 |
| Total |  | 31758 | 2413 | 546 | 1704 | 36421 |

**Supplementary Table 2:**
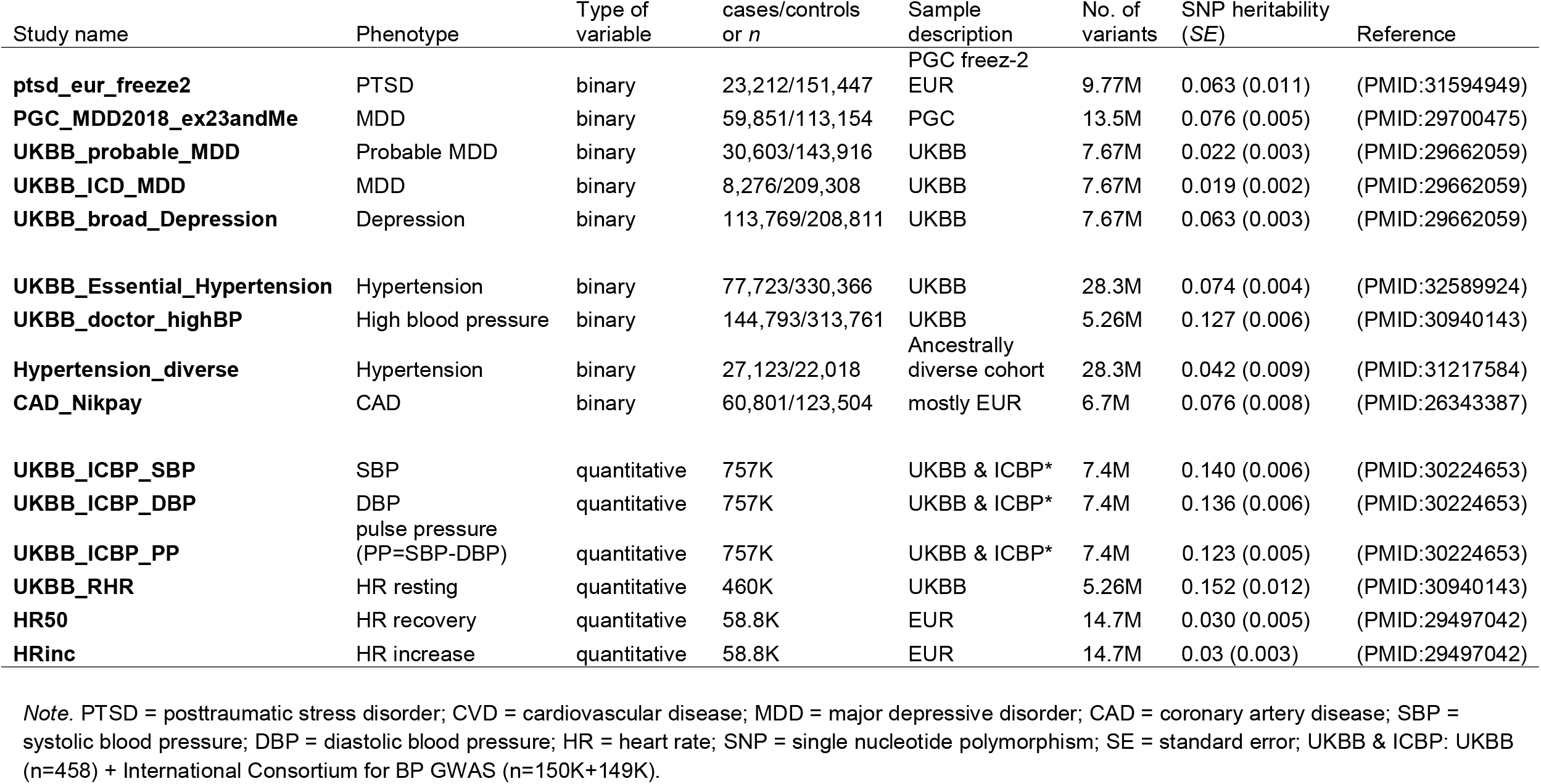
List of publicly available summary statistics used in the current study.

**Supplementary Table 3:**
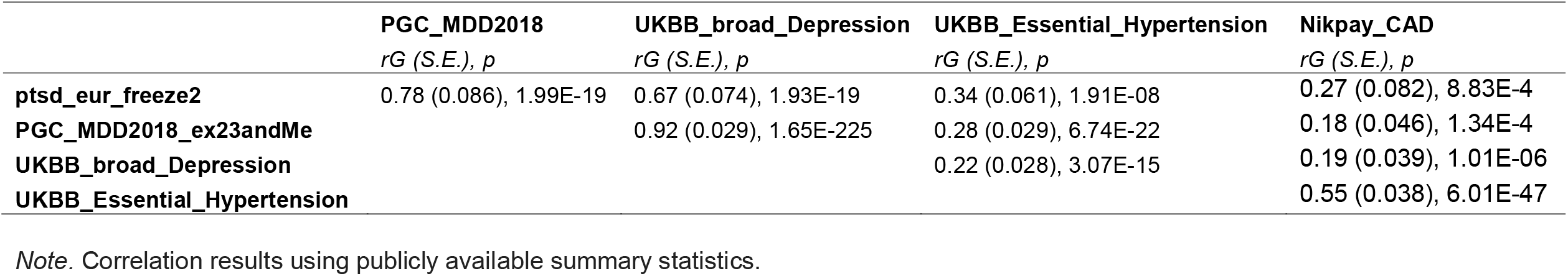
Genetic correlations among PTSD/MDD and CVD (hypertension/CAD).

**Supplementary Table 4:**
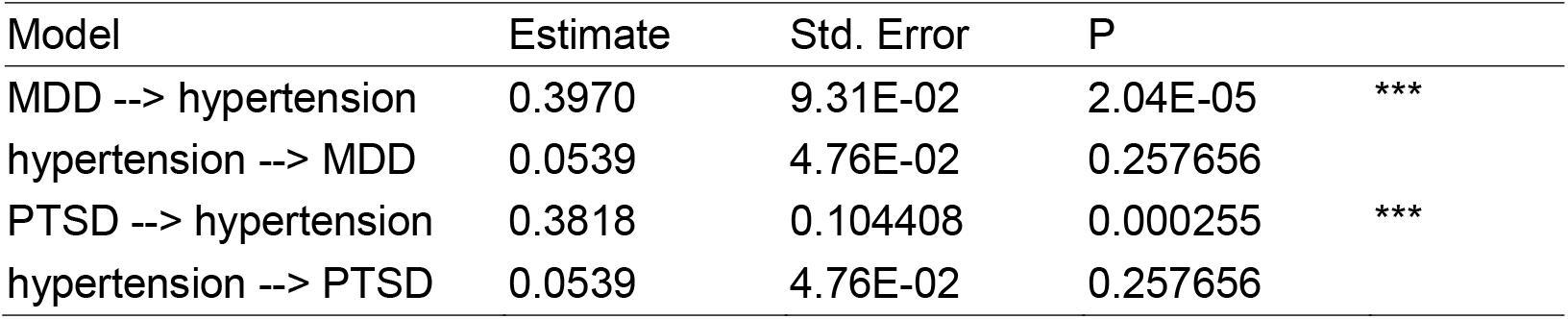
Mendelian randomization results suggest causal pathway from PTSD and MDD to hypertension

**Supplementary Figure 1:**
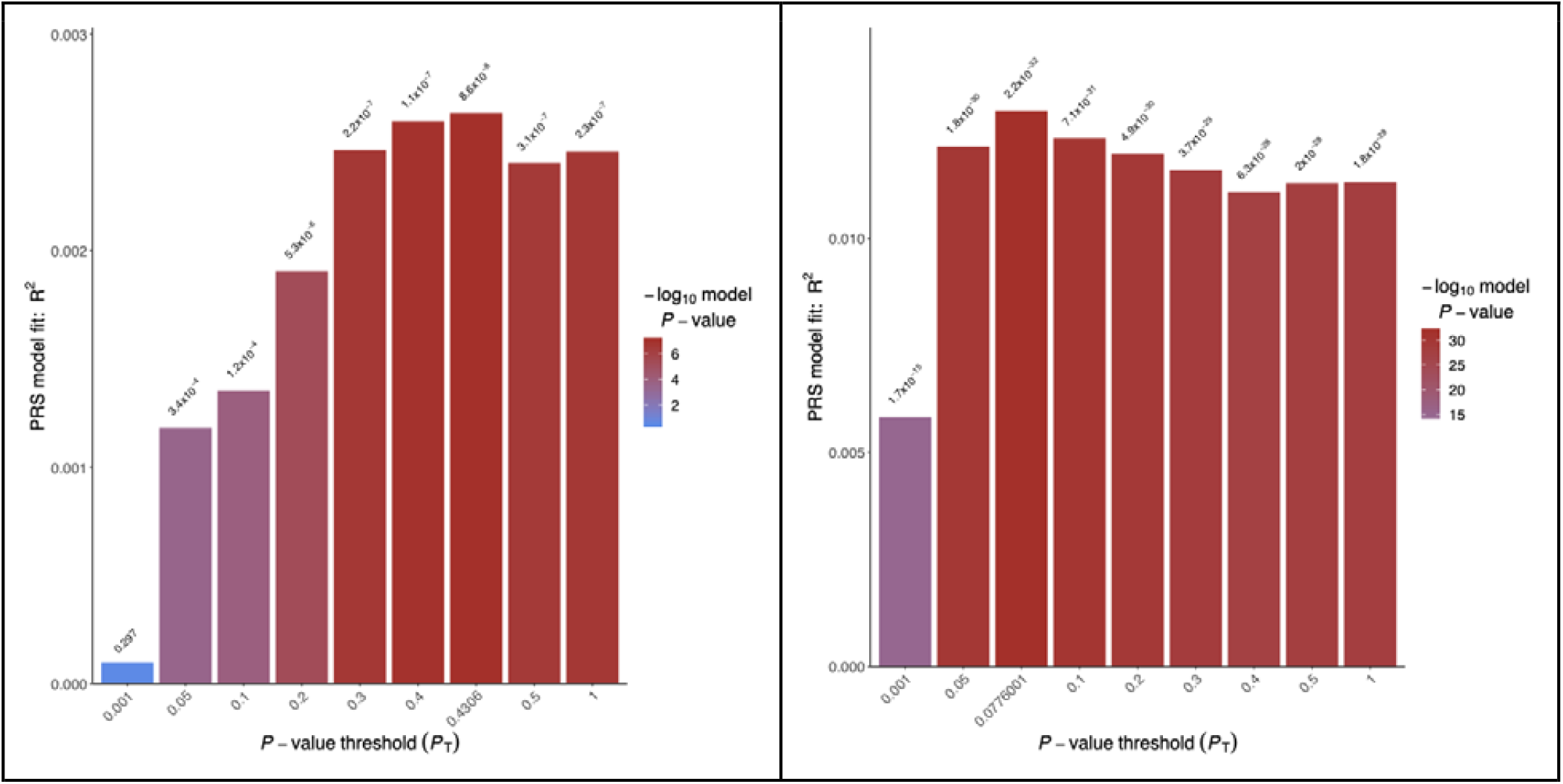
Thresholding step with MTAG summary statistics for PTSD polygenic risk score (PRS).

**Supplementary Figure 2:**
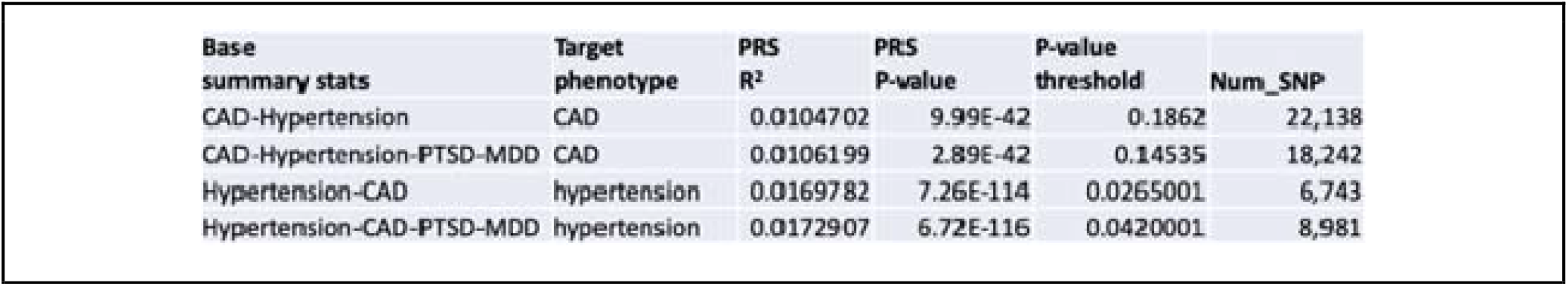
Improvement in the prediction of coronary artery disease (CAD) and hypertension using PTSD and MDD summary statistics.

**Supplementary Figure 3:**
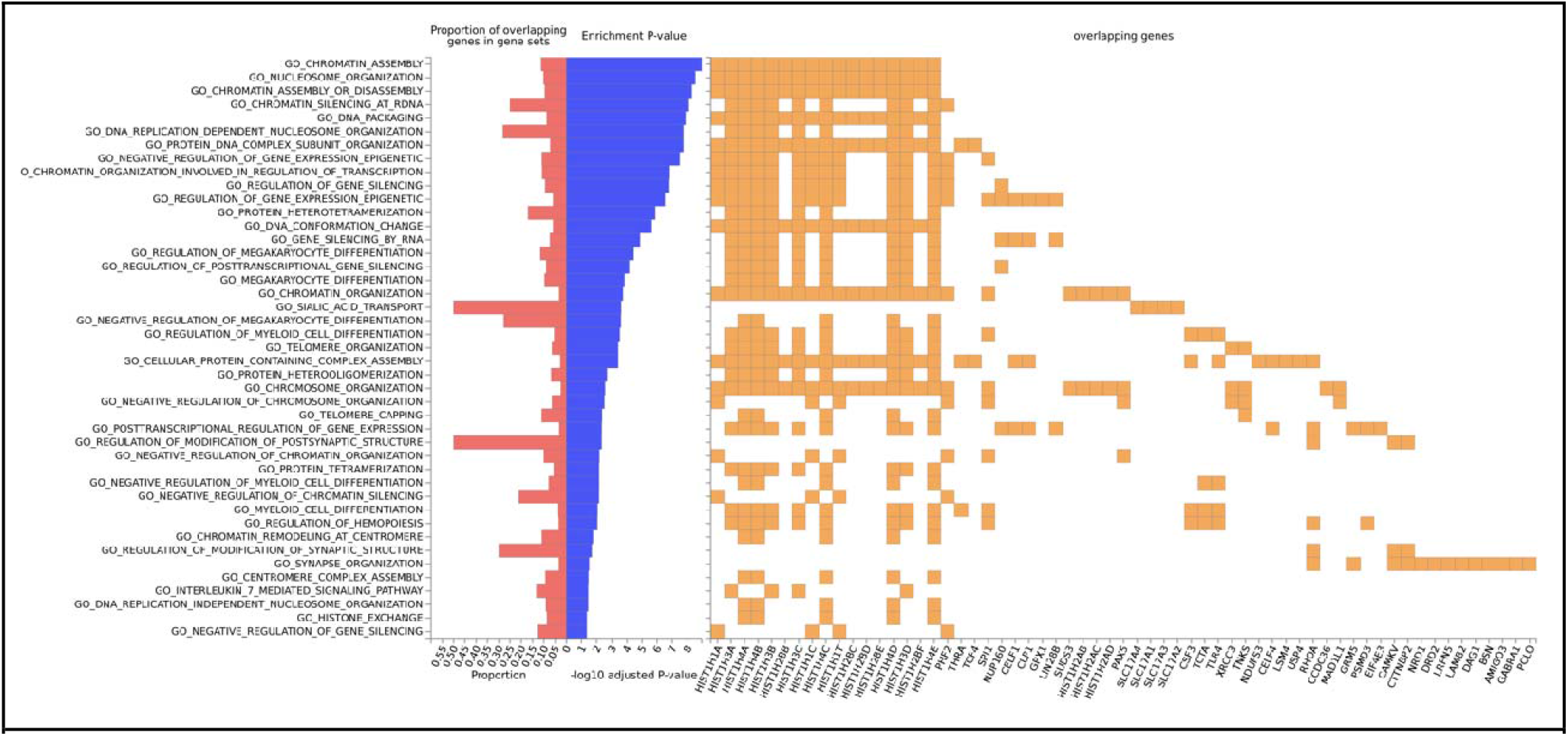
Genomic risk loci included in pathway analysis.

**Supplementary Figure 4:**
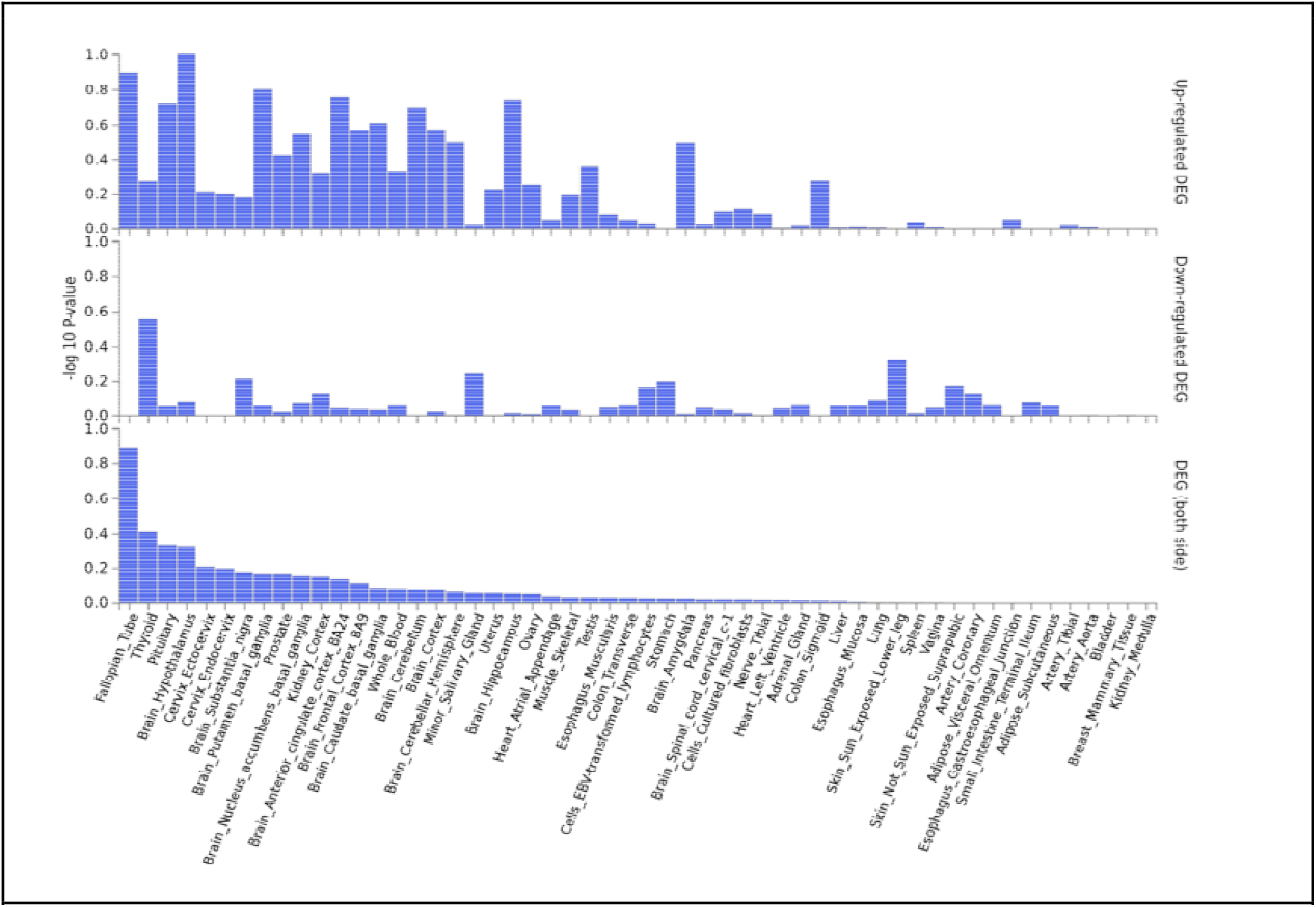
Tissue specificity of pathway analysis.

