## Supplementary File for "Leveraging large-scale genetics of PTSD and cardiovascular disease demonstrates robust shared risk and improves risk prediction accuracy"

**Supplementary Material**

**Phenotype data**

Diagnostic data was extracted from the MGBB and ICD-10 codes and curated disease populations were used to determine diagnosis.^31,32^ Curated Disease Populations are collected by a validated phenotype algorithm with a positive predictive value of 0.90. These algorithms rely on coded diagnosis as well as natural language processing terms extracted from clinical narratives. Predictive features for each characterization were identified using an automated feature extraction protocol, which identifies comorbidities, symptoms, and medications. Concepts were screened based on frequency in patient clinical notes. Billing diagnosis and prescriptions were also included in algorithms.^31,32^ Patients with lifetime history of congestive heart failure, coronary artery disease, hypertension, ischemic stroke, and depression as determined by Curated Disease Population status were considered positive for these traits in our dataset. Curated Disease Populations were not available for PTSD or myocardial infarction.

**Curated Disease Population Algorithms**

Model definitions with features and betas: Below are the feature weights of the final hypertension phenotype algorithms. The weights below are used to derive a predicted probability of hypertension or no hypertension for every Biobank participant.

| **Feature_ID** | **Beta (weight)** | **Feature Description** |
| --- | --- | --- |
| (Intercept) | 0.247 | Model Intercept (beta 0) |
| HTN_COD_DX_Hypertension | 0.793 | Count of coded diagnosis of hypertension |
| HTN_COD_MED_Antihypertensives | 0.627 | Count of prescriptions for an antihypertensive |
| HTN_COD_MED_Aceinhibitor | 0.476 | Count of prescriptions for an ACE inhibitor |
| max_bmi_new | -0.323 | Maximum BMI (if missing, impute to mean sample BMI of 30) |
| patient_dxenct | -0.586 | Total number of visits with a coded diagnosis |

Below are the feature weights of the final CAD phenotype algorithms. The weights below can be used to derive a predicted probability of CAD or no CAD for any Biobank participant.

| **Feature_ID** |  | **Beta (weight)** | **Feature Description** |
| --- | --- | --- | --- |
| (Intercept) |  | 5.288948213 | Model Intercept (beta 0) |
| patient_dxenct |  | -3.121364389 | Number of encounters with ICD-9 codes |
| CAD_COD_DX_IschemicHeartDisease |  | 1.934743408 | Coded mentions of a Ischemic Heart Disease diagnosis |
| CAD_NLP_alcohol |  | 0.16107732 | Hitex non-negated NLP mentions of UMLS CUI C0001962: alcohol |
| CAD_NLP_angioplasty |  | 0.022473675 | Hitex non-negated NLP mentions of UMLS CUI C0162577: angioplasty |
| CAD_NLP_antiplateletagents |  | 0.503346441 | Hitex non-negated NLP mentions of UMLS CUI C0085826: antiplatelet agents |
| CAD_NLP_coronaryarterybypassgrafting |  | 1.054659295 | Hitex non-negated NLP mentions of UMLS CUI C0010055: coronary artery bypass grafting |
| CAD_NLP_coronaryatherosclerosis |  | -1.194440371 | Hitex non-negated NLP mentions of UMLS CUI C0010054: coronary atherosclerosis |
| CAD_NLP_coronaryheartdisease |  | 2.588093909 | Hitex non-negated NLP mentions of UMLS CUI C0010068: coronary heart disease |
| CAD_NLP_creatinine |  | -0.342506235 | Hitex non-negated NLP mentions of UMLS CUI C0010294: creatinine |
| CAD_NLP_electrocardiogram |  | -1.62020437 | Hitex non-negated NLP mentions of UMLS CUI C0013798: electrocardiogram |
| CAD_NLP_ischemia |  | 1.037838709 | Hitex non-negated NLP mentions of UMLS CUI C0022116: ischemia |
| CAD_NLP_ischemiccardiomyopathy |  | -0.106484107 | Hitex non-negated NLP mentions of UMLS CUI C0349782: ischemic cardiomyopathy |
| CAD_NLP_myocardialinfarction |  | 0.230624677 | Hitex non-negated NLP mentions of UMLS CUI C0027051: myocardial infarction |
| CAD_NLP_nitroglycerin |  | 1.769966347 | Hitex non-negated NLP mentions of UMLS CUI C0017887: nitroglycerin |
| CAD_NLP_plateletaggregationinhibitors |  | 0.433529361 | Hitex non-negated NLP mentions of UMLS CUI C0032177: platelet aggregation inhibitors |

Below are the feature weights of the final depression phenotype algorithms. The weights below are used to derive a predicted probability of depression or no depression for every Biobank participant.

| **Feature_ID** | **Beta (weight)** | **Feature Description** |
| --- | --- | --- |
| (Intercept) | -2.539 | Model Intercept (beta 0) |
| Depression_COD_DX_Depression | 1.973 | Count of coded diagnosis of depression or MDD |
| Depression_COD_DX_Mentalhealthdisorders | 0.175 | Count of coded diagnosis of any mental health disorder |
| Depression_COD_DX_Bipolardisorder | -0.813 | Count of coded diagnosis of bipolar disorder |
| Depression_COD_MED_Antidepressants | 1.096 | Count of prescriptions for an antidepressant |
| Depression_COD_MED_Antipsychotics | -0.395 | Count of prescriptions for an antipsychotic |
| Depression_COD_MED_Anticonvulsants | -0.323 | Count of prescriptions for an anticonvulsant |
| patient_dxenct | -0.515 | Total number of visits with a coded diagnosis |

Below are the feature weights of the final CHF phenotype algorithms. The weights below can be used to derive a predicted probability of CHF or no CHF for any Biobank participant.

| **Feature_ID** | **Beta (weight)** | **Feature Description** |
| --- | --- | --- |
| (Intercept) | -0.85309914 | Model Intercept (beta 0) |
| patient_dxenct | -0.790872064 | Number of encounters with an ICD-9 code |
| CHF_COD_DX_CHF | 0.299480034 | Count of CHF diagnoses |
| CHF_NLP_heartfailure | 0.36842715 | Hitex non-negated NLP mentions of UMLS CUI C0018802: heart failure |
| CHF_NLP_ischemiccardiomyopathy | 0.39455684 | Hitex non-negated NLP mentions of UMLS CUI C0349782: ischemic cardiomyopathy |
| CHF_NLP_loopdiuretics | 0.650788804 | Hitex non-negated NLP mentions of UMLS CUI C0354100: loop diuretics |

Below are the feature weights of the final Ischemic stroke phenotype algorithms. The weights below are used to derive a predicted probability of ISTR for every Biobank participant.

| **Feature_ID** | **Beta (weight)** | **Feature Description** |
| --- | --- | --- |
| (Intercept) | 2.640 | Model Intercept (beta 0) |
| Stroke_COD_DX_IschemicStroke | 1.205 | Count of coded diagnosis of ischemic stroke |
| Stroke_COD_DX_HemorrhagicStroke | -0.507 | Count of coded diagnosis of hemorrhagic stroke |
| Stroke_NLP_brainattack | 0.828 | Count of non-negated NLP mentions of brain attack |
| Stroke_NLP_transientischaemicattack | -0.137 | Count of non-negated NLP mentions of transient ischemic attack |
| patient_dxenct | -1.480 | Total number of visits with a coded diagnosis |

| Estimate | Std. Error | P | Significance |
| --- | --- | --- | --- |
| 0.428 | 0.095 | 5.97E-06 | *** |

Therefore, we can infer that MDD onset is linked to hypertension diagnosis. We implemented a similar MR procedure for three other models. The results are shown in the table below.

| Model | beta | Std. Error | *p* |
| --- | --- | --- | --- |
| MDD --> hypertension | 0.428 | 0.095 | 5.97e-06 |
| MDD --> CAD | 0.466 | 0.141 | 9.22e-04 |
| PTSD --> hypertension | 0.201 | 0.044 | 5.97e-06 |
| PTSD --> CAD | 0.219 | 0.066 | 9.22e-04 |

| Model | beta | Std. Error | *p* |
| --- | --- | --- | --- |
| hypertension --> MDD | 0.054 | 0.048 | 0.258 |
| hypertension --> PTSD | 0.144 | 0.068 | 0.0345 |
| CAD --> MDD | 0.157 | 0.075 | 0.0373 |
| CAD --> PTSD | 0.310 | 0.108 | 0.0041 |

As shown, our analysis provides support for a causal link from psychiatric disorders to cardiovascular illnesses, but not the other way around.

Note that all analysis is only for EUR sub-population of the MGBB dataset. In the future, similar analysis on sub-populations (based on ancestry, gender and age) needs to be conducted. All analysis is done on R statistical software.

**CRP analyses**

We investigated genetic level correlations between the two groups of phenotypes and the largest publicly available CRP GWAS summary statistics (PMID: 33462484). As expected, CRP level in the blood is genetically correlated with both group of phenotypes. The main result is summarized below.

| Summary statistics | r_g_ | P |
| --- | --- | --- |
| Hypertension_meta | 0.3183 | 5.98E-16 |
| UKBB_Essential_Hypertension | 0.3206 | 6.06E-16 |
| CAD_meta | 0.2254 | 1.04E-07 |
| UKBB_broad_Depression | 0.2326 | 4.63E-07 |
| PGC_MDD2018_ex23andMe | 0.1691 | 2.70E-05 |
| MDD_meta | 0.1622 | 5.46E-05 |
| PTSD_meta | 0.2211 | 0.000138 |
| pts_eur_freeze2 | 0.1859 | 0.001913 |

**Supplementary Tables and Figures:**

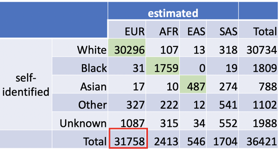

Supplementary Table 1: Self-identified and genetically predicted ancestry composition. Genetic analysis was done with those participants predicted to have EUR ancestry.

Supplementary Table 2: List of publicly available summary statistics used in the current study.

| Study name | Phenotype | Type of variable | cases/controls or *n* | Sample description | No. of variants | SNP heritability (*SE*) | Reference |
| --- | --- | --- | --- | --- | --- | --- | --- |
| **ptsd_eur_freeze2** | PTSD | binary | 23,212/151,447 | PGC freez-2 EUR | 9.77M | 0.063 (0.011) | (PMID:31594949) |
| **PGC_MDD2018_ex23andMe** | MDD | binary | 59,851/113,154 | PGC | 13.5M | 0.076 (0.005) | (PMID:29700475) |
| **UKBB_probable_MDD** | Probable MDD | binary | 30,603/143,916 | UKBB | 7.67M | 0.022 (0.003) | (PMID:29662059) |
| **UKBB_ICD_MDD** | MDD | binary | 8,276/209,308 | UKBB | 7.67M | 0.019 (0.002) | (PMID:29662059) |
| **UKBB_broad_Depression** | Depression | binary | 113,769/208,811 | UKBB | 7.67M | 0.063 (0.003) | (PMID:29662059) |
| **UKBB_Essential_Hypertension** | Hypertension | binary | 77,723/330,366 | UKBB | 28.3M | 0.074 (0.004) | (PMID:32589924) |
| **UKBB_doctor_highBP** | High blood pressure | binary | 144,793/313,761 | UKBB | 5.26M | 0.127 (0.006) | (PMID:30940143) |
| **Hypertension_diverse** | Hypertension | binary | 27,123/22,018 | Ancestrally diverse cohort | 28.3M | 0.042 (0.009) | (PMID:31217584) |
| **CAD_Nikpay** | CAD | binary | 60,801/123,504 | mostly EUR | 6.7M | 0.076 (0.008) | (PMID:26343387) |
| **UKBB_ICBP_SBP** | SBP | quantitative | 757K | UKBB & ICBP* | 7.4M | 0.140 (0.006) | (PMID:30224653) |
| **UKBB_ICBP_DBP** | DBP | quantitative | 757K | UKBB & ICBP* | 7.4M | 0.136 (0.006) | (PMID:30224653) |
| **UKBB_ICBP_PP** | pulse pressure (PP=SBP-DBP) | quantitative | 757K | UKBB & ICBP* | 7.4M | 0.123 (0.005) | (PMID:30224653) |
| **UKBB_RHR** | HR resting | quantitative | 460K | UKBB | 5.26M | 0.152 (0.012) | (PMID:30940143) |
| **HR50** | HR recovery | quantitative | 58.8K | EUR | 14.7M | 0.030 (0.005) | (PMID:29497042) |
| **HRinc** | HR increase | quantitative | 58.8K | EUR | 14.7M | 0.03 (0.003) | (PMID:29497042) |

*Note.* PTSD = posttraumatic stress disorder; CVD = cardiovascular disease; MDD = major depressive disorder; CAD = coronary artery disease; SBP = systolic blood pressure; DBP = diastolic blood pressure; HR = heart rate; SNP = single nucleotide polymorphism; SE = standard error; UKBB & ICBP: UKBB (n=458) + International Consortium for BP GWAS (n=150K+149K).

Supplementary Table 3: Genetic correlations among PTSD/MDD and CVD (hypertension/CAD).

|  | **PGC_MDD2018** | **UKBB_broad_Depression** | **UKBB_Essential_Hypertension** | **Nikpay_CAD** |
| --- | --- | --- | --- | --- |
|  | *rG (S.E.), p* | *rG (S.E.), p* | *rG (S.E.), p* | *rG (S.E.), p* |
| **ptsd_eur_freeze2** | 0.78 (0.086), 1.99E-19 | 0.67 (0.074), 1.93E-19 | 0.34 (0.061), 1.91E-08 | 0.27 (0.082), 8.83E-4 |
| **PGC_MDD2018_ex23andMe** |  | 0.92 (0.029), 1.65E-225 | 0.28 (0.029), 6.74E-22 | 0.18 (0.046), 1.34E-4 |
| **UKBB_broad_Depression** |  |  | 0.22 (0.028), 3.07E-15 | 0.19 (0.039), 1.01E-06 |
| **UKBB_Essential_Hypertension** |  |  |  | 0.55 (0.038), 6.01E-47 |

*Note.* Correlation results using publicly available summary statistics.

Supplementary Table 4: Mendelian randomization results suggest causal pathway from PTSD and MDD to hypertension

| Model | Estimate | Std. Error | P |  |
| --- | --- | --- | --- | --- |
| MDD --> hypertension | 0.3970 | 9.31E-02 | 2.04E-05 | *** |
| hypertension --> MDD | 0.0539 | 4.76E-02 | 0.257656 |  |
| PTSD --> hypertension | 0.3818 | 0.104408 | 0.000255 | *** |
| hypertension --> PTSD | 0.0539 | 4.76E-02 | 0.257656 |  |

| 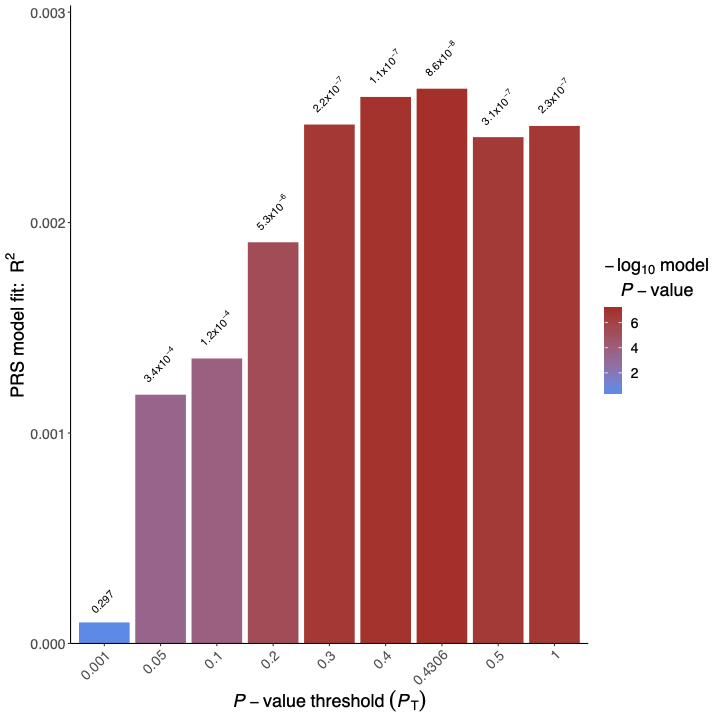 | 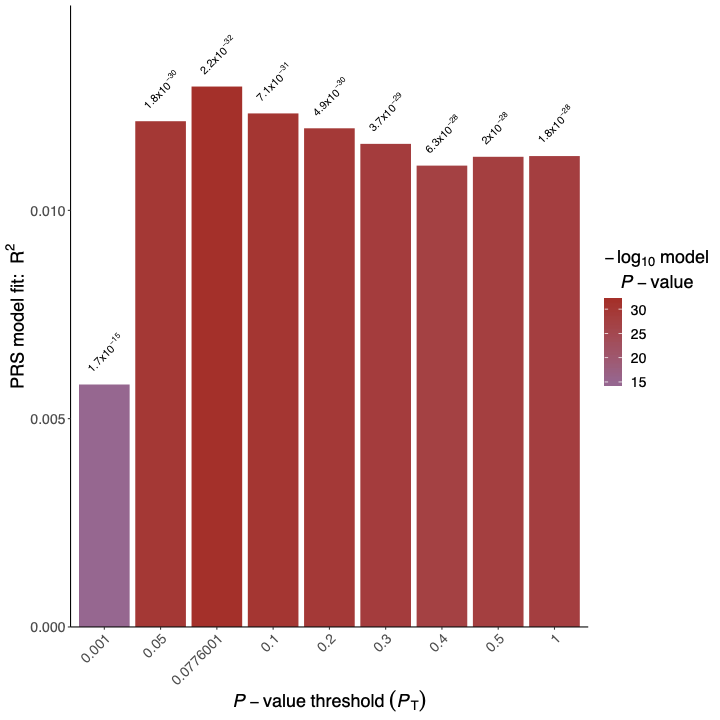 |
| --- | --- |
| Supplementary Figure 1: Thresholding step with MTAG summary statistics for PTSD polygenic risk score (PRS). | |

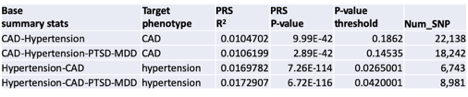

Supplementary Figure 2: Improvement in the prediction of coronary artery disease (CAD) and hypertension using PTSD and MDD summary statistics.

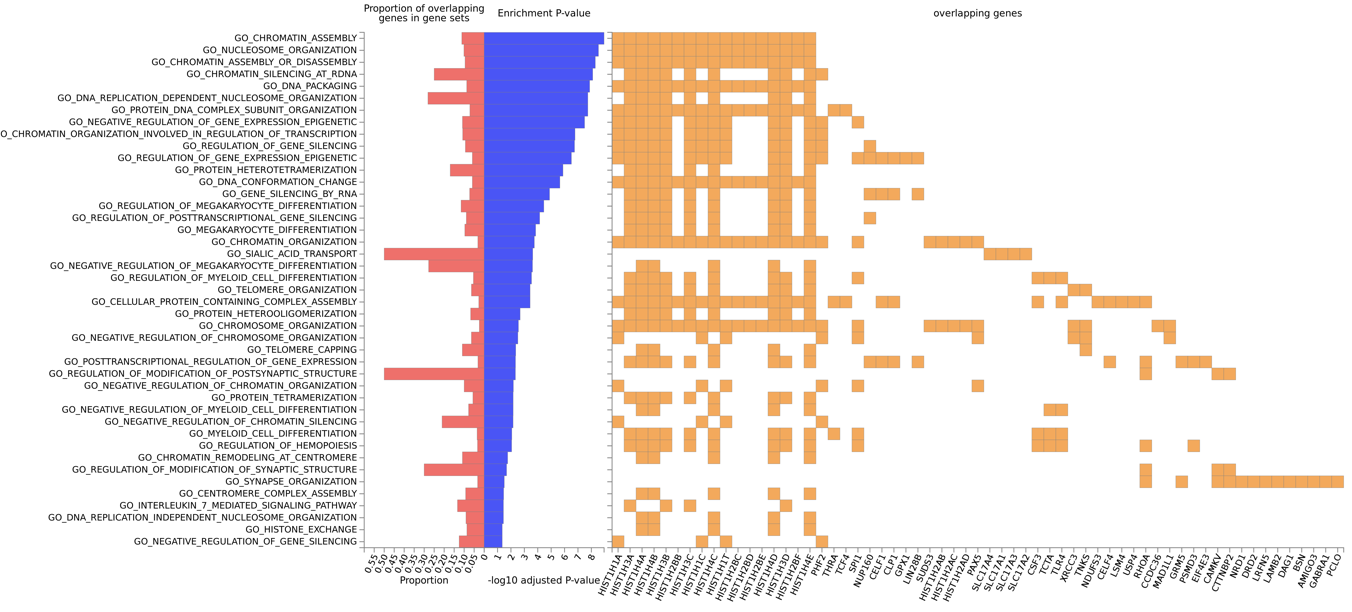

Supplementary Figure 3: Genomic risk loci included in pathway analysis.

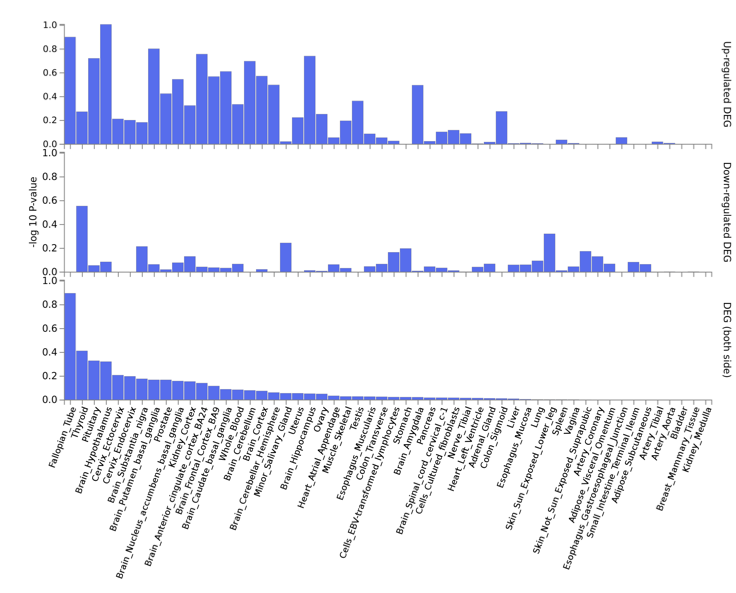

Supplementary Figure 4: Tissue specificity of pathway analysis.
