## Supplementary Figures for "Leveraging large-scale genetics of PTSD and cardiovascular disease demonstrates robust shared risk and improves risk prediction accuracy"

Supplementary Figure 5: MGBB_CAD_manhattan
*Note*. Manhattan plot for coronary artery disease (CAD); MGBB = MassGeneral Brigham Biobank.

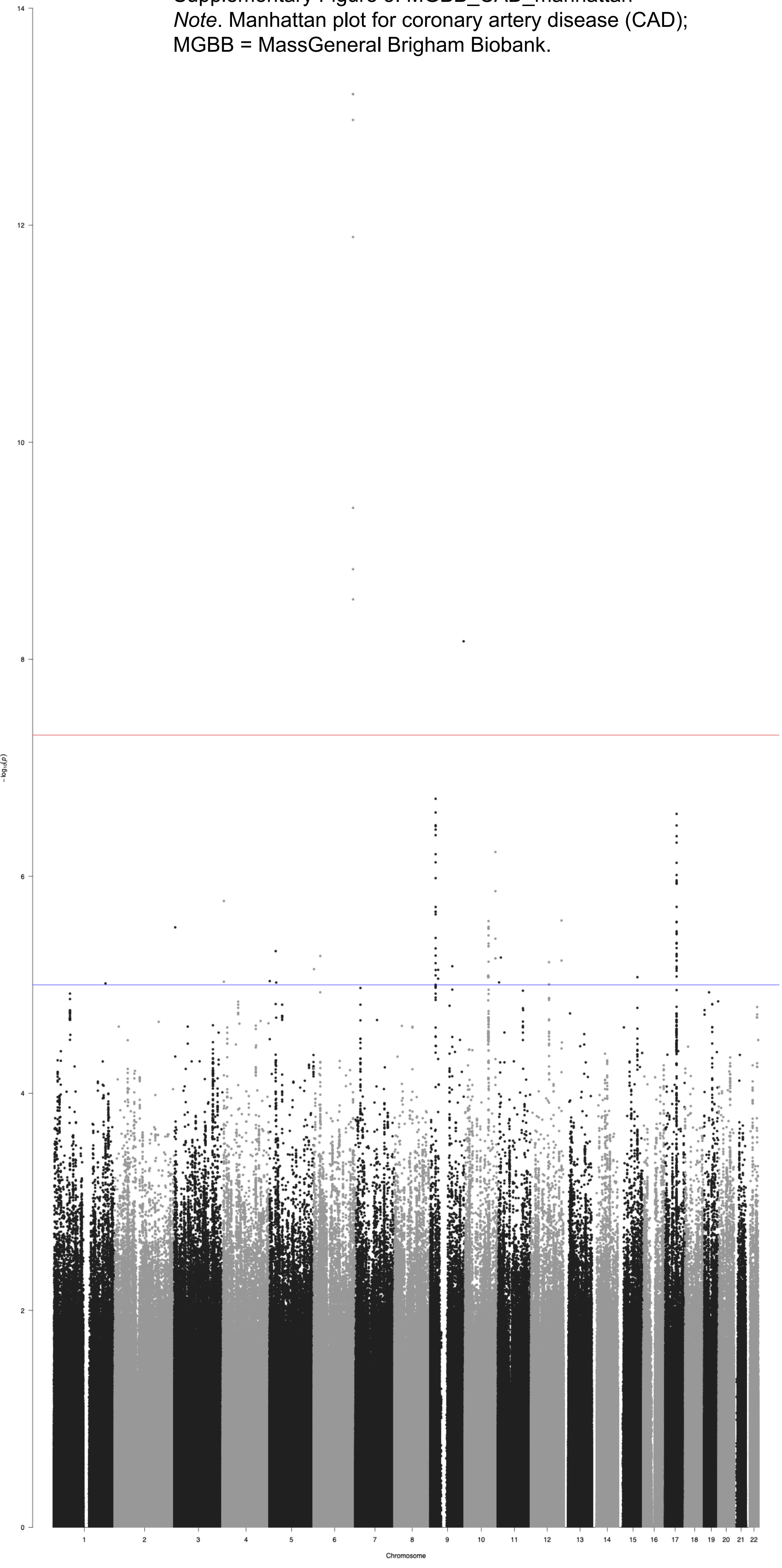

Supplementary Figure 6: MGBB_CAD_qq
*Note*. QQ plot for coronary artery disease (CAD);
MGBB = MassGeneral Brigham Biobank.

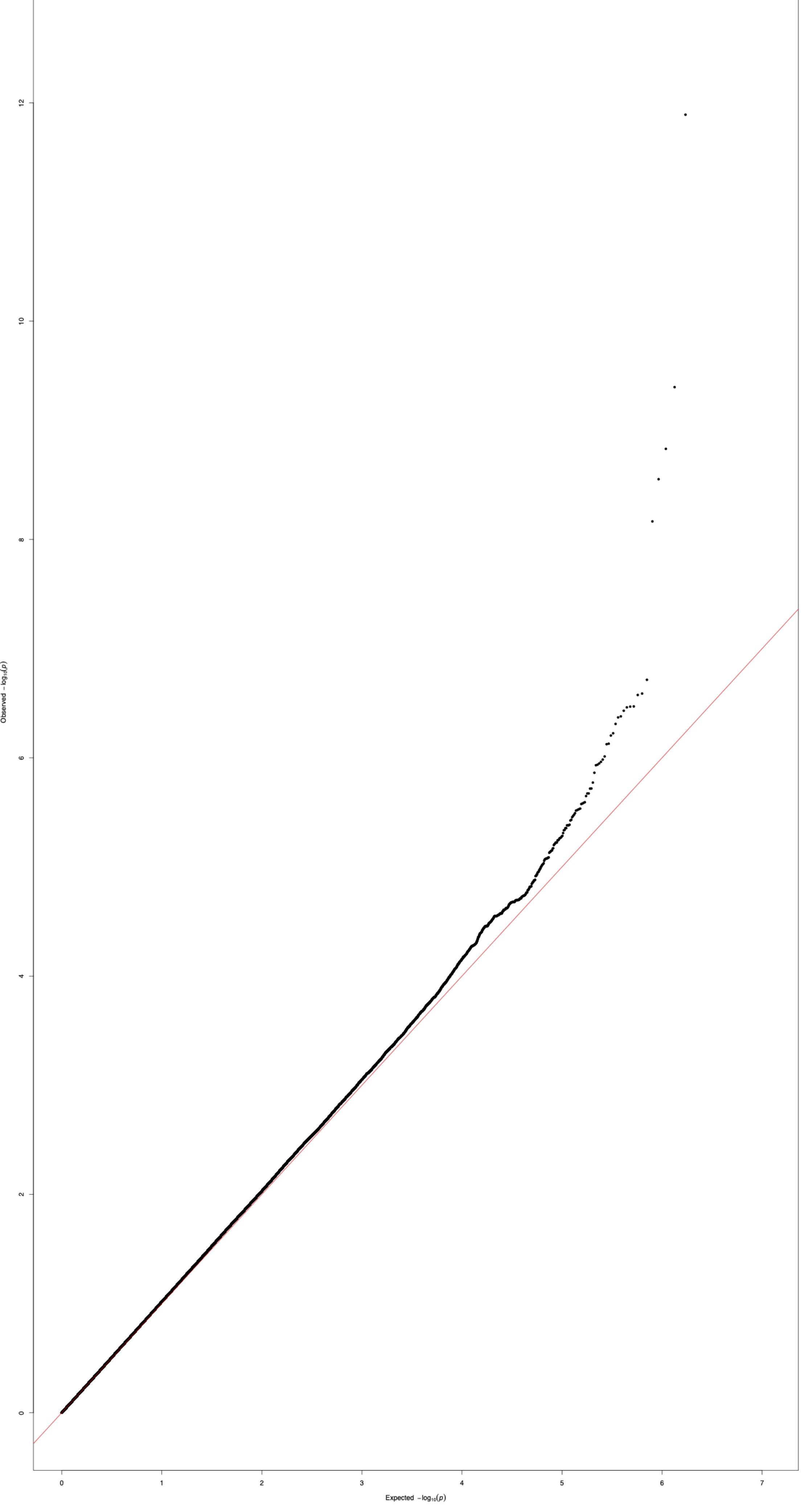

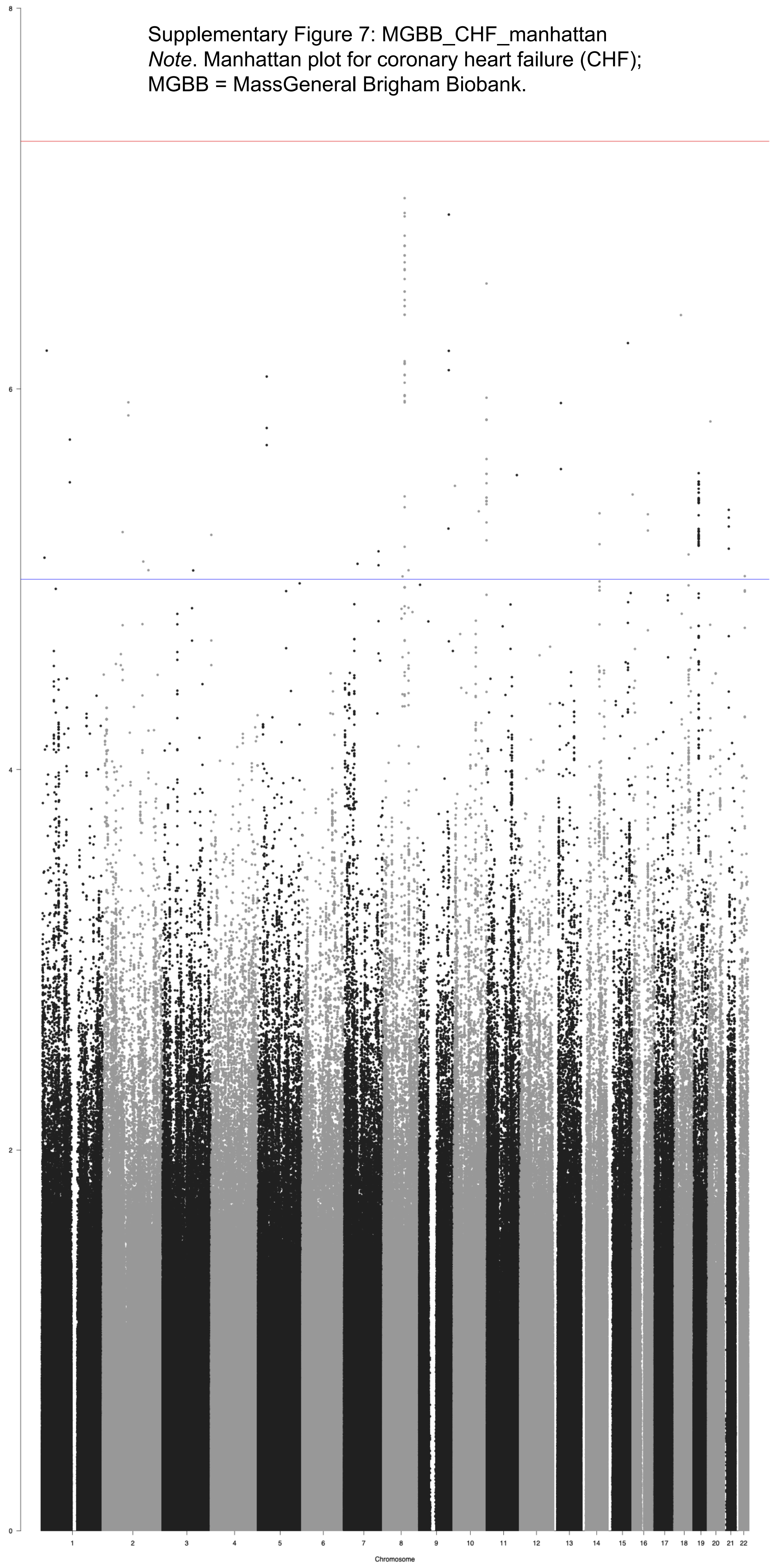


Supplementary Figure 7: MGBB_CHF_manhattan
*Note*. Manhattan plot for coronary heart failure (CHF);
MGBB = MassGeneral Brigham Biobank.

Supplementary Figure 8: MGBB_CHF_qq
*Note*. QQ plot for coronary heart failure (CHF);
MGBB = MassGeneral Brigham Biobank.

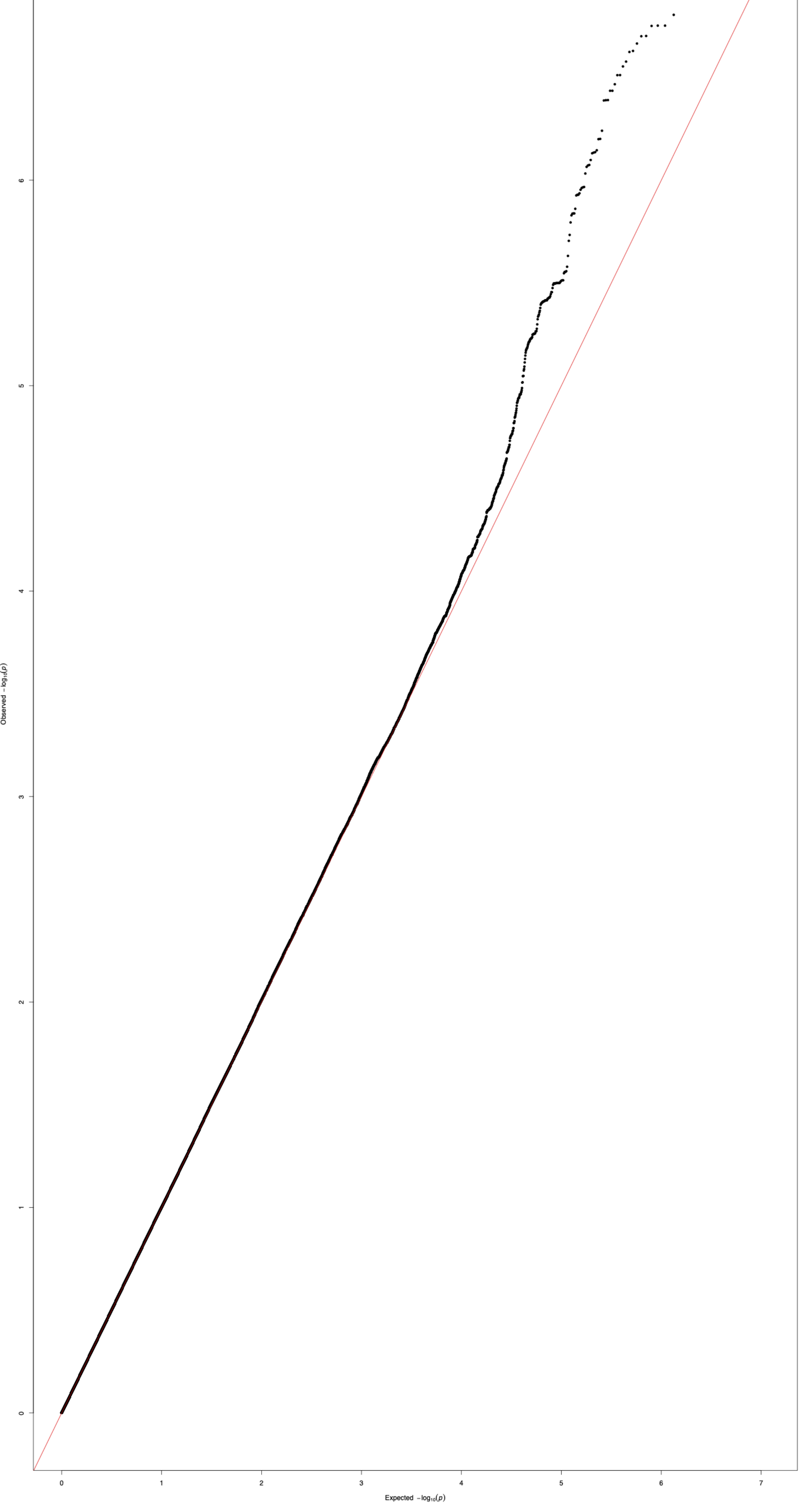

Supplementary Figure 9: MGBB_DiastolicAvg_manhattan
*Note*. Manhattan plot for Average Diastolic Blood Pressure; MGBB = MassGeneral Brigham Biobank.

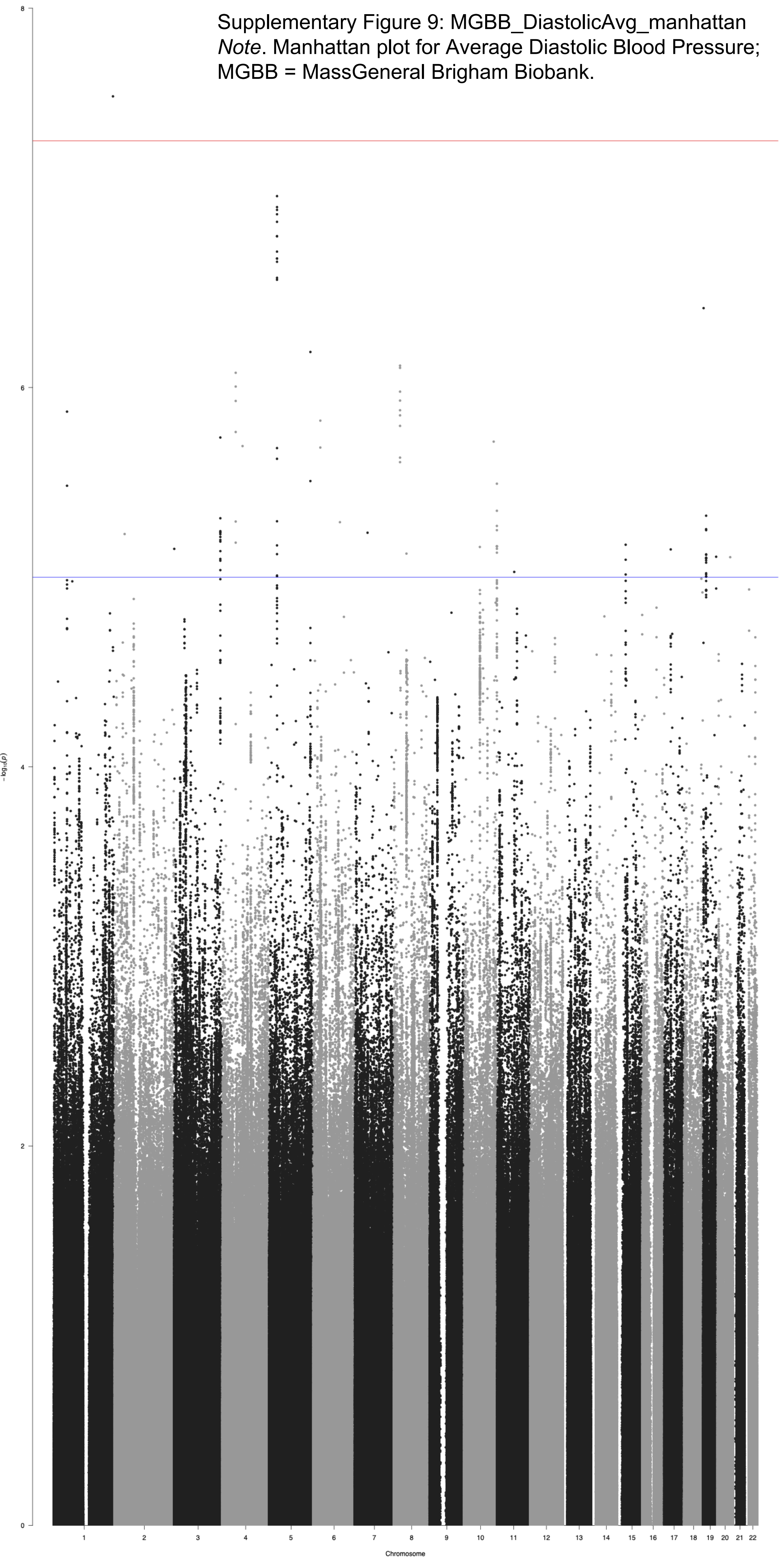

Supplementary Figure 10: MGBB_DiastolicAvg_qq
*Note*. QQ plot for Average Diastolic Blood Pressure;
MGBB = MassGeneral Brigham Biobank.

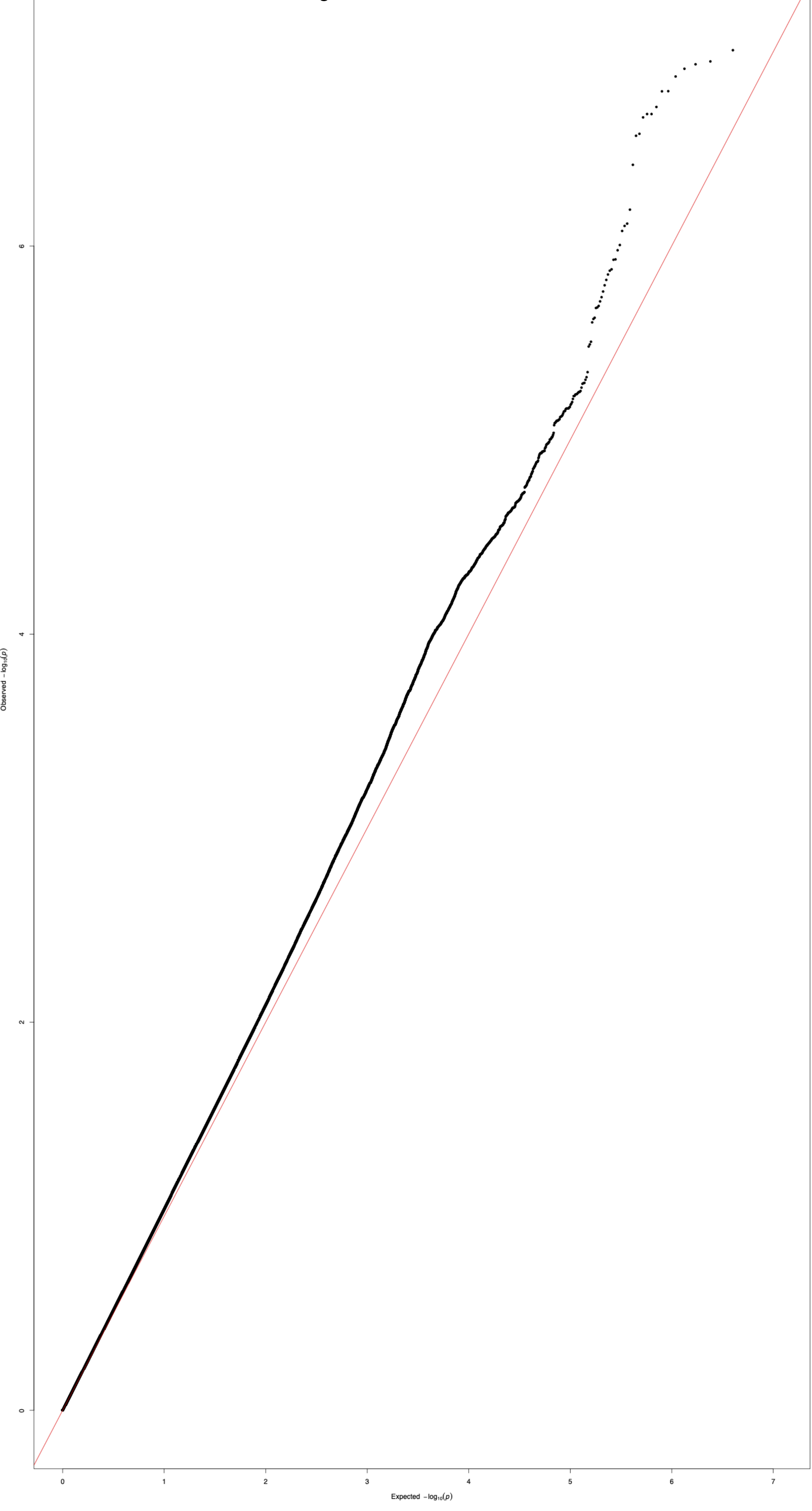

Supplementary Figure 11: MGBB_Hypertension_manhattan
*Note*. Manhattan plot for Hypertension; MGBB = MassGeneral Brigham Biobank.

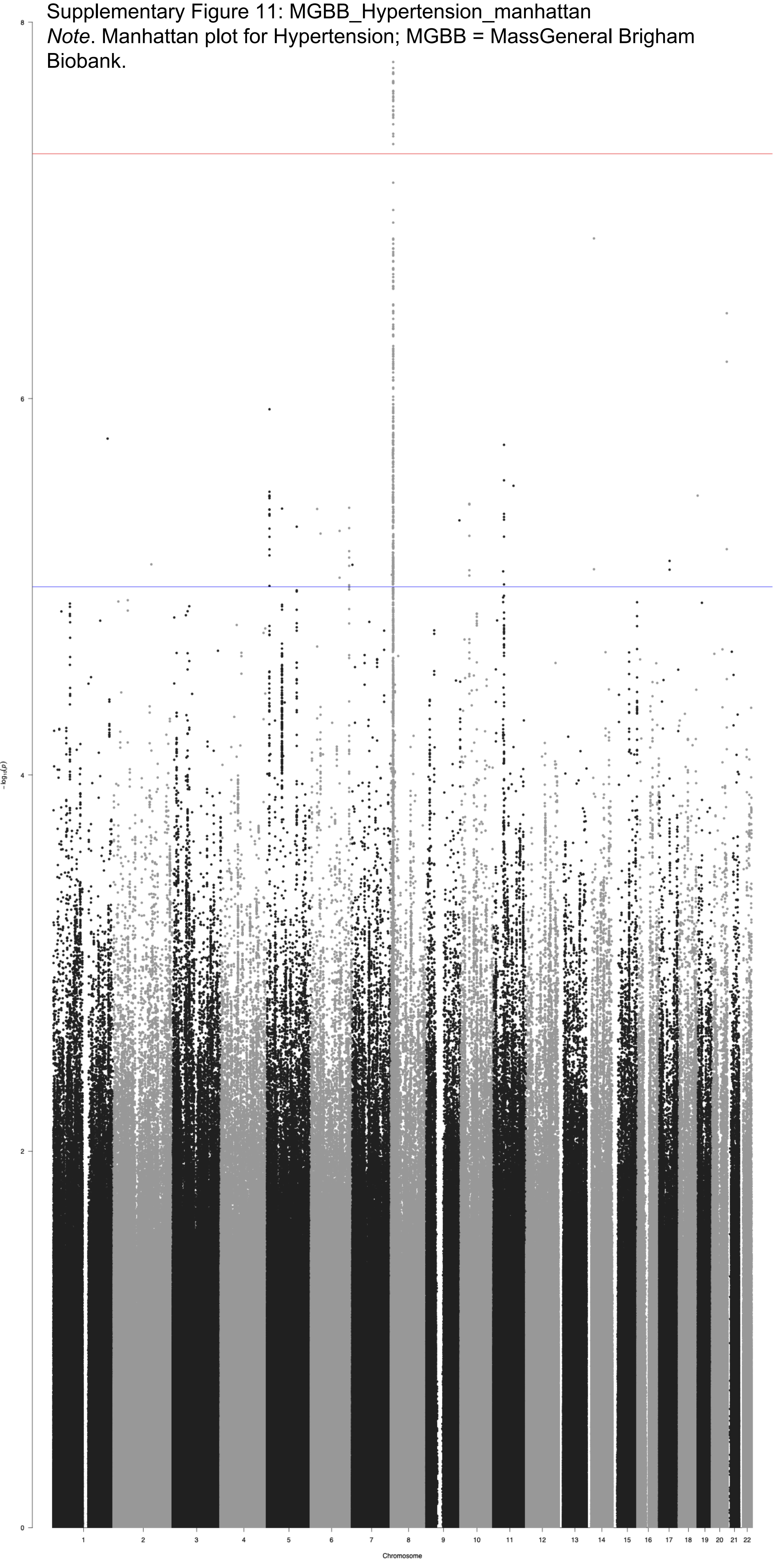

Supplementary Figure 12: MGBB_Hypertension_qq
*Note*. QQ plot for Hypertension; MGBB = MassGeneral Brigham Biobank.

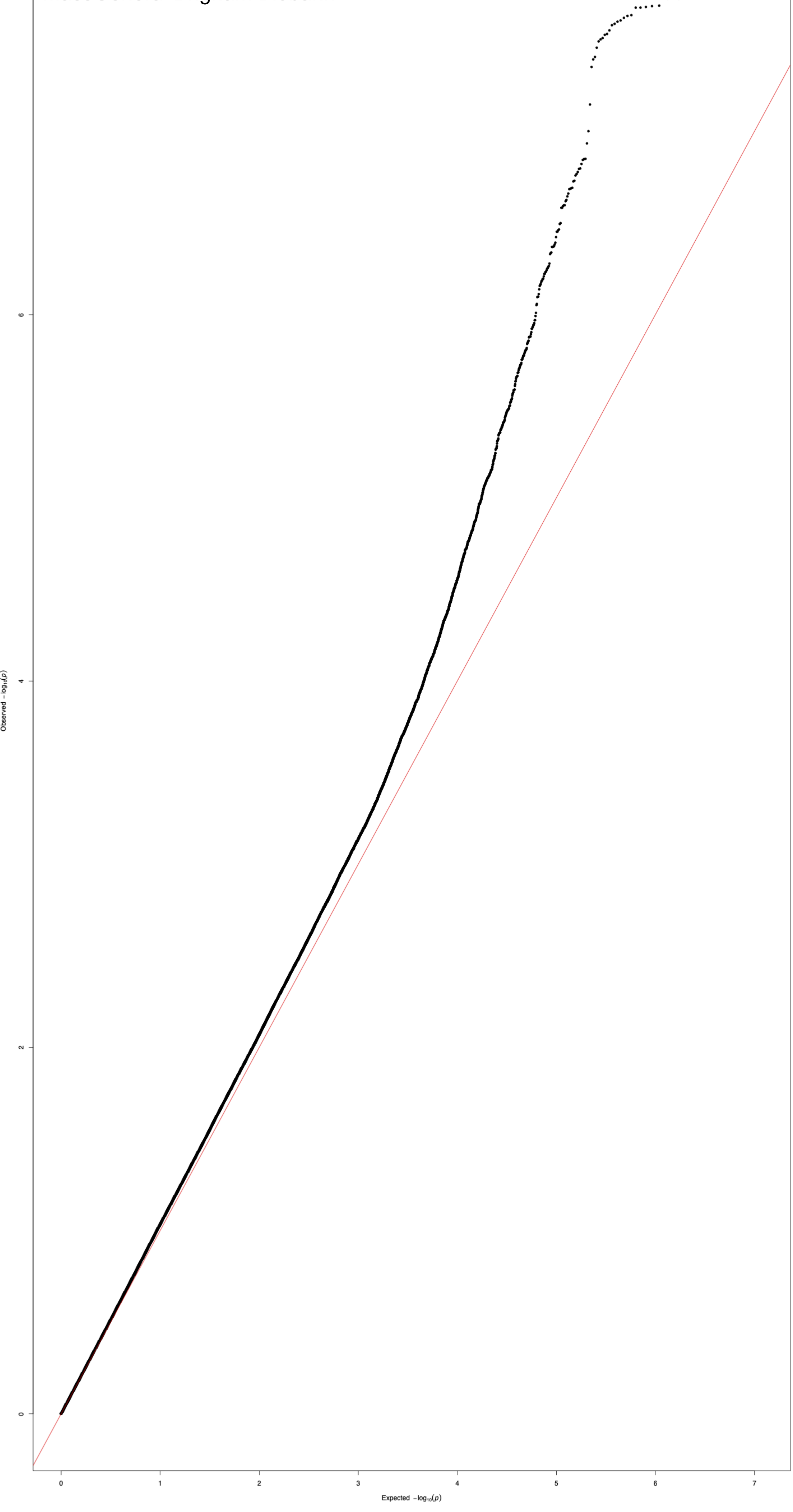

Supplementary Figure 13: MGBB_MDD_manhattan

*Note*. Manhattan plot for Major Depressive Disorder (MDD); MGBB = MassGeneral Brigham Biobank.

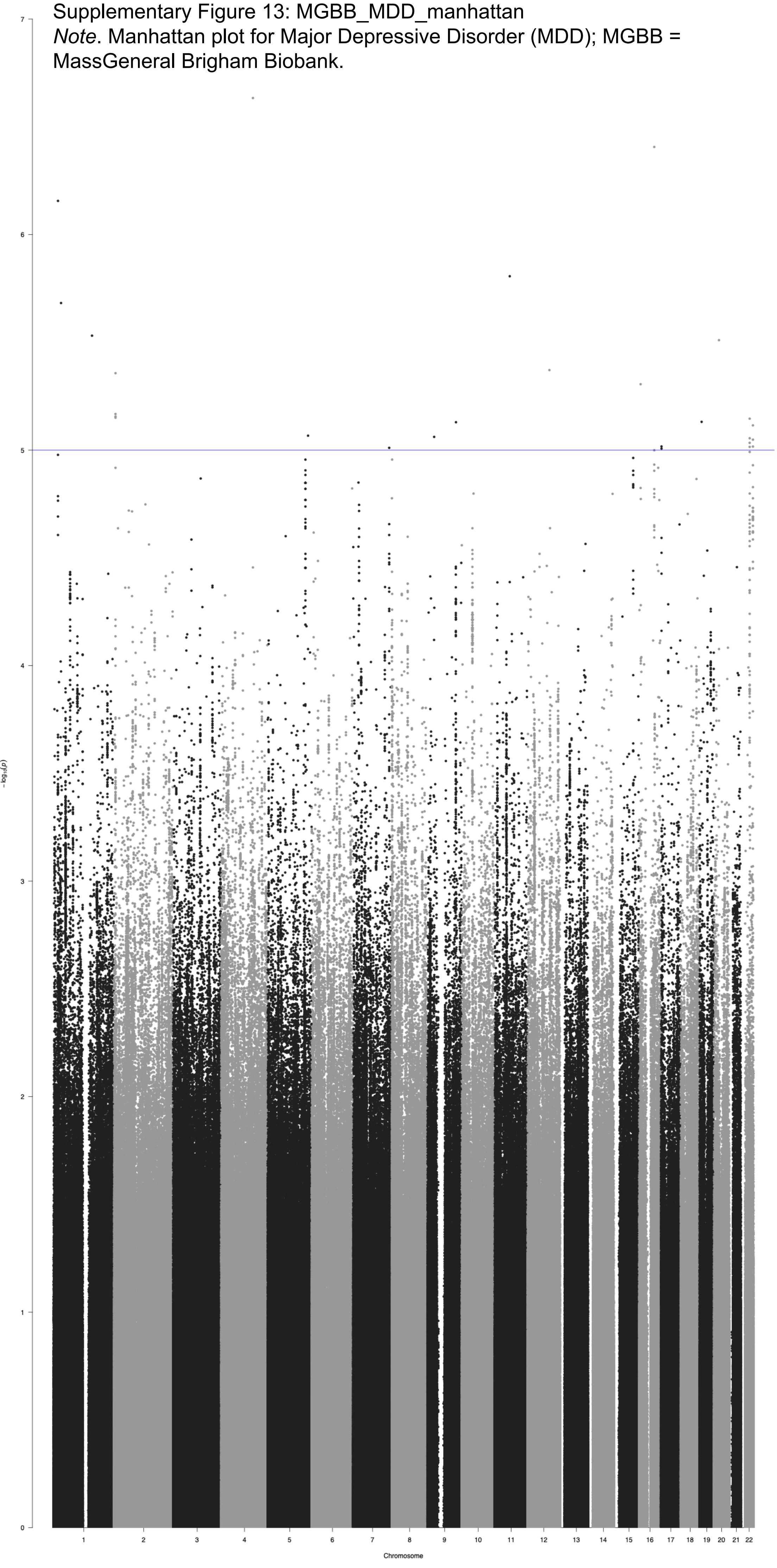

Supplementary Figure 14: MGBB_MDD_qq
*Note*. QQ plot for Major Depressive Disorder (MDD);
MGBB = MassGeneral Brigham Biobank.

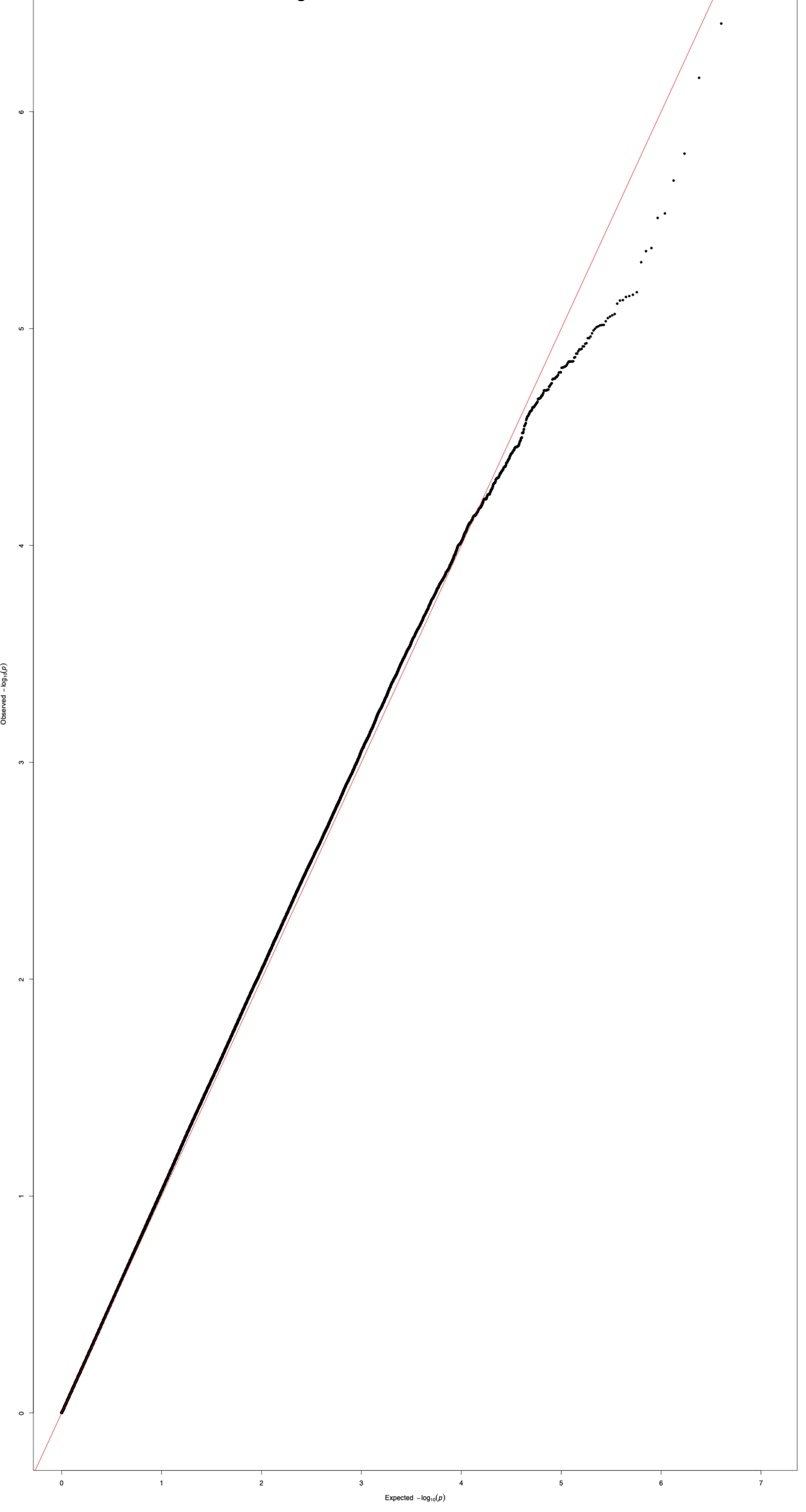

Supplementary Figure 15: MGBB_MI_manhattan
*Note*. Manhattan plot for Myocardial Infarction (MI); MGBB = MassGeneral Brigham Biobank.

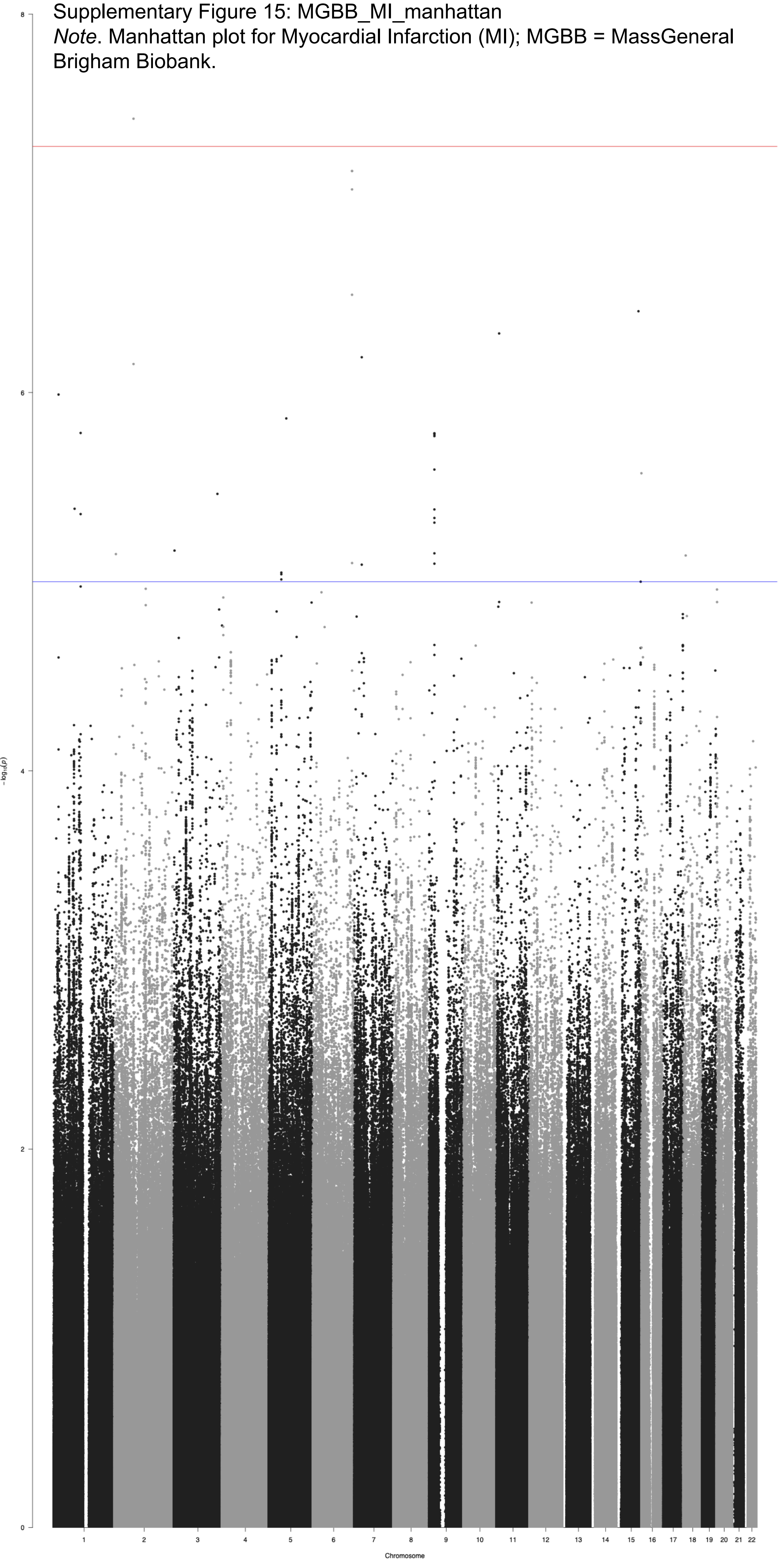

Supplementary Figure 16: MGBB_MI_qq
*Note*. QQ plot for Myocardial Infarction (MI); MGBB = MassGeneral Brigham Biobank.

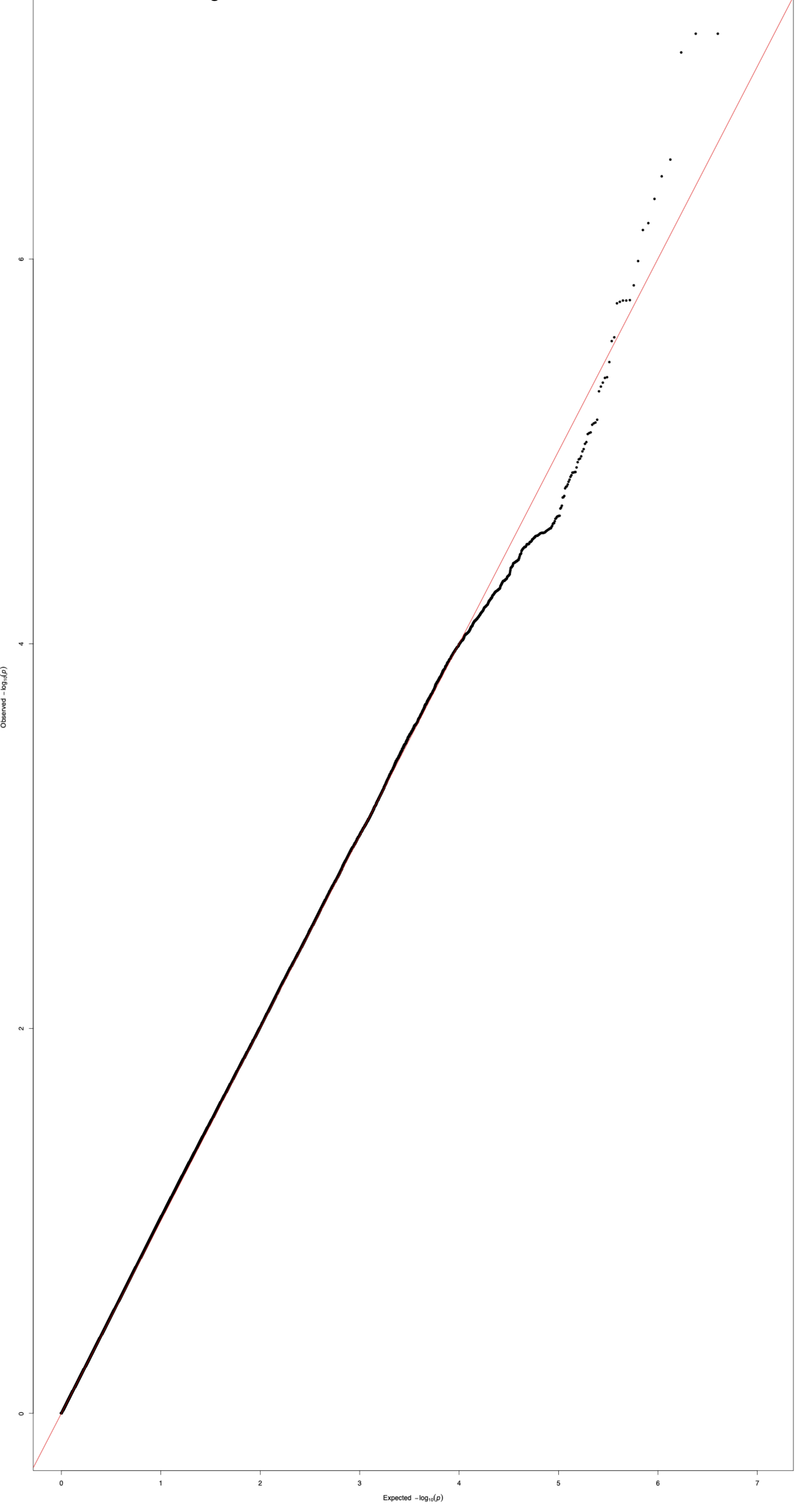

Supplementary Figure 17: MGBB_PTSD_manhattan

*Note*. Manhattan plot for PTSD; MGBB = MassGeneral Brigham Biobank.

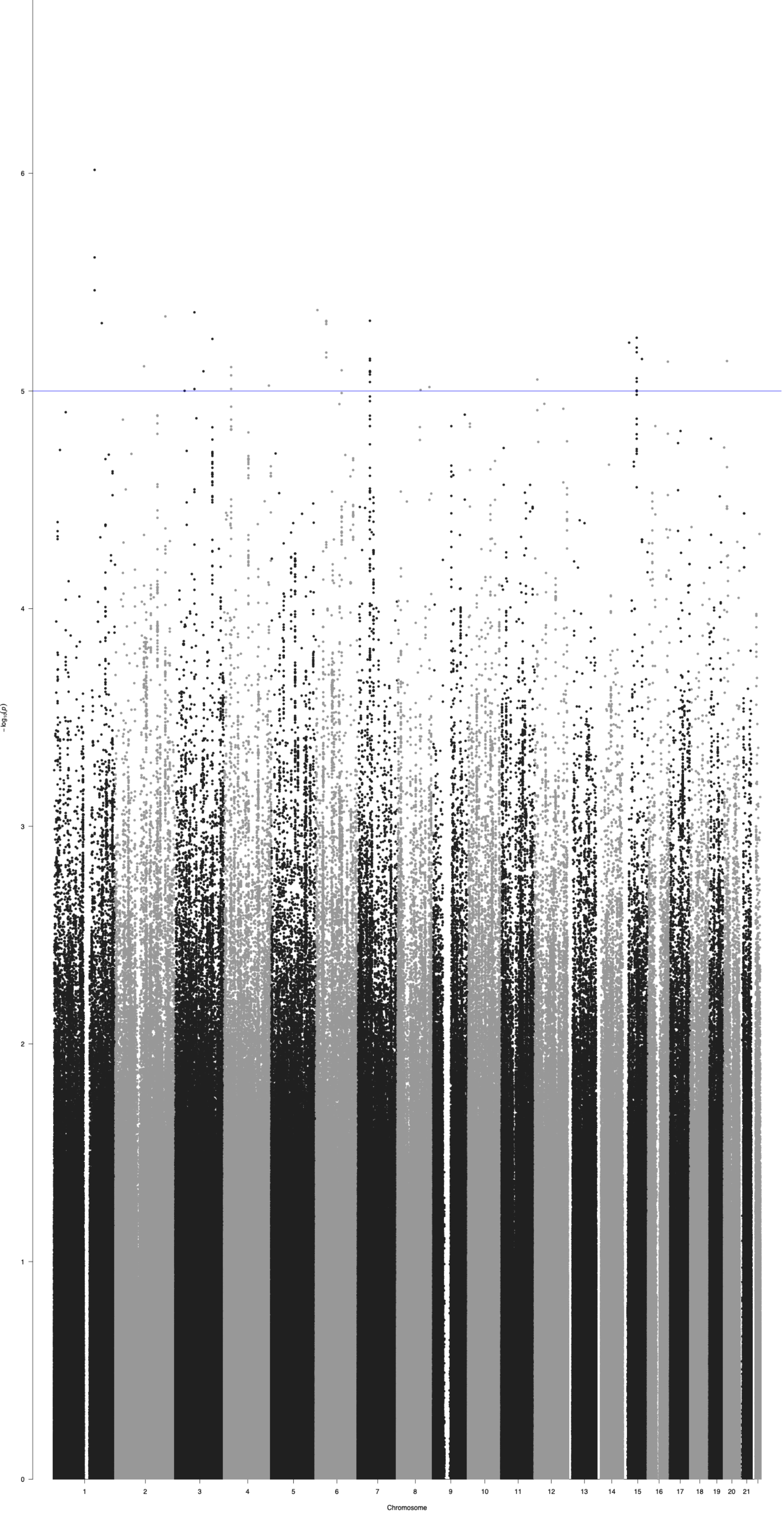

Supplementary Figure 18: MGBB_PTSD_qq
*Note*. QQ plot for PTSD; MGBB = MassGeneral Brigham Biobank.

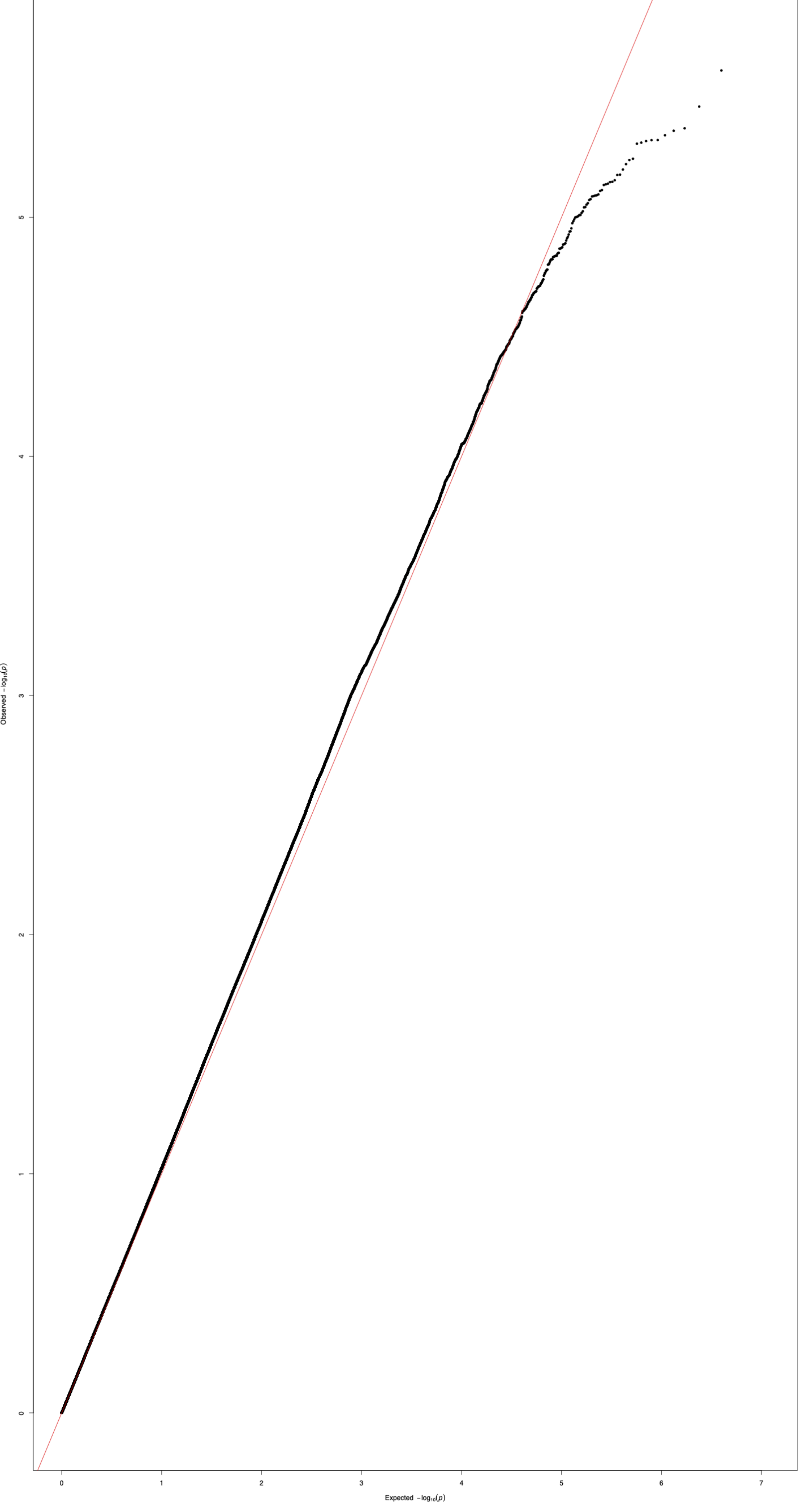

Supplementary Figure 19: MGBB_PulseAvg_manhattan
*Note*. Manhattan plot for Average Pulse; MGBB = MassGeneral Brigham Biobank.

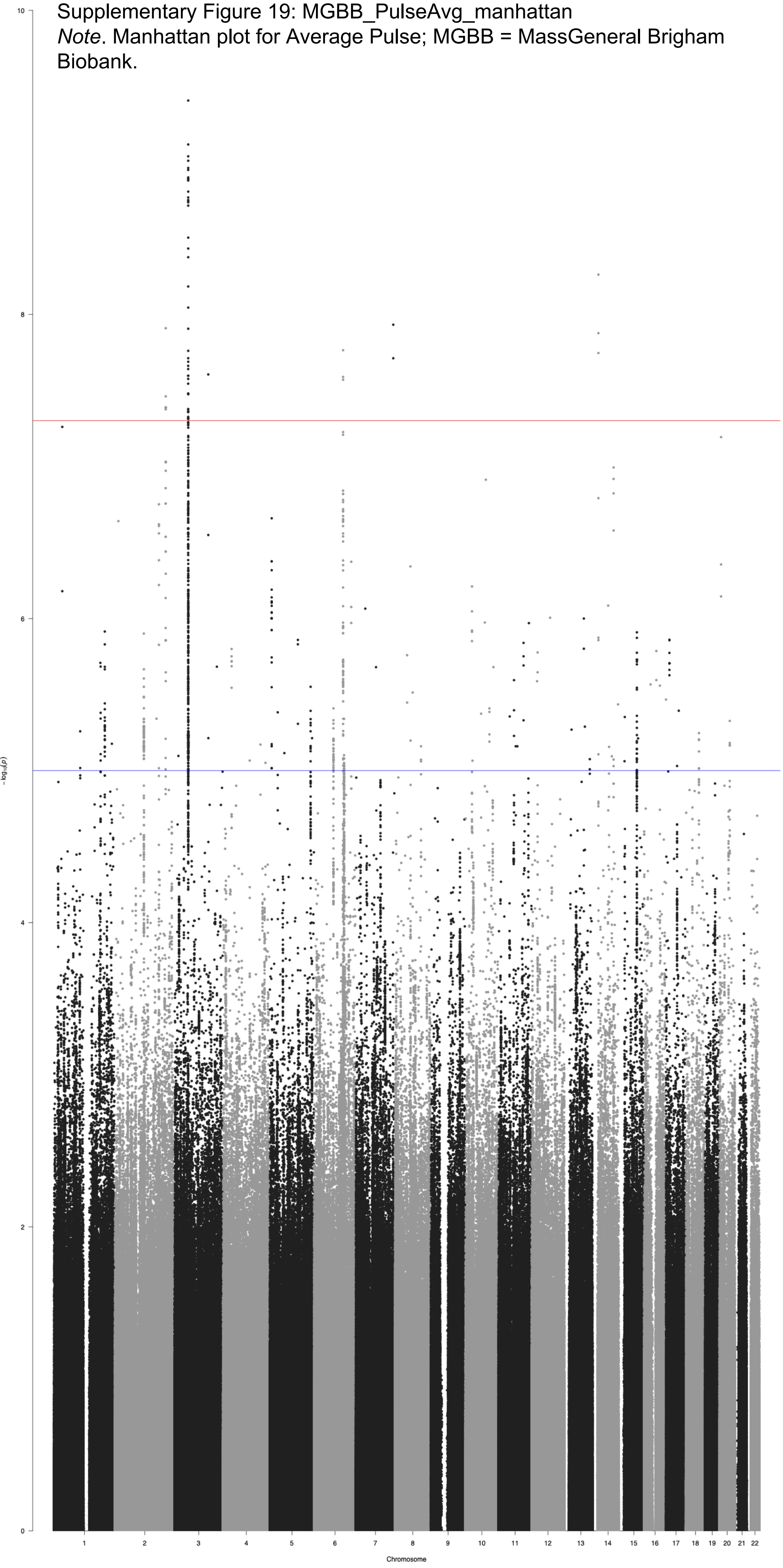

Supplementary Figure 20: MGBB_PulseAvg_qq

*Note*. QQ plot for Average Pulse; MGBB = MassGeneral Brigham Biobank.

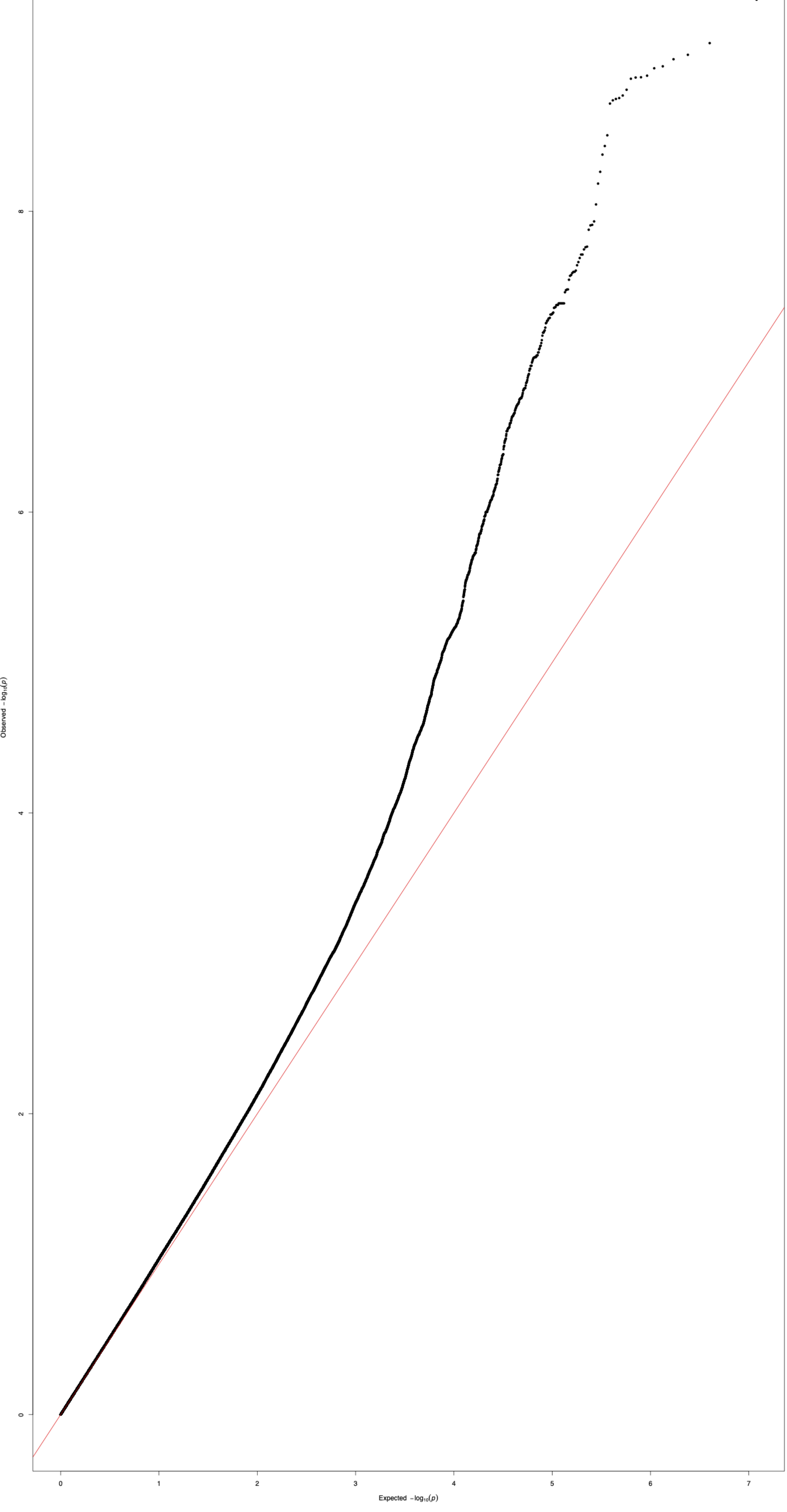

Supplementary Figure 21: MGBB_SystolicAvg_manhattan

*Note*. Manhattan plot for Average Systolic Blood Pressure; MGBB = MassGeneral Brigham Biobank.

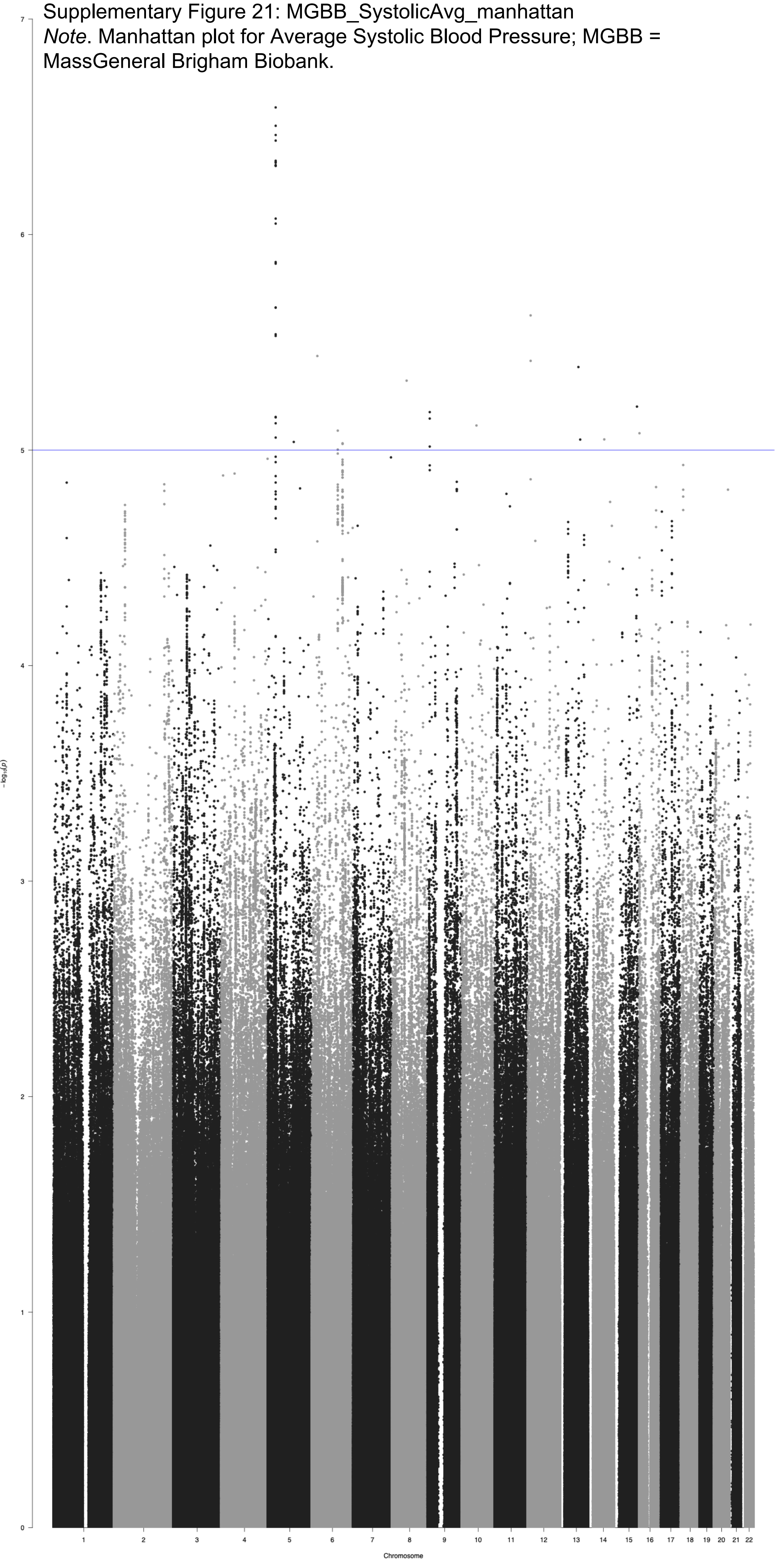

Supplementary Figure 22: MGBB_SystolicAvg_qq
*Note*. QQ plot for Average Systolic Blood Pressure; MGBB = MassGeneral Brigham Biobank.

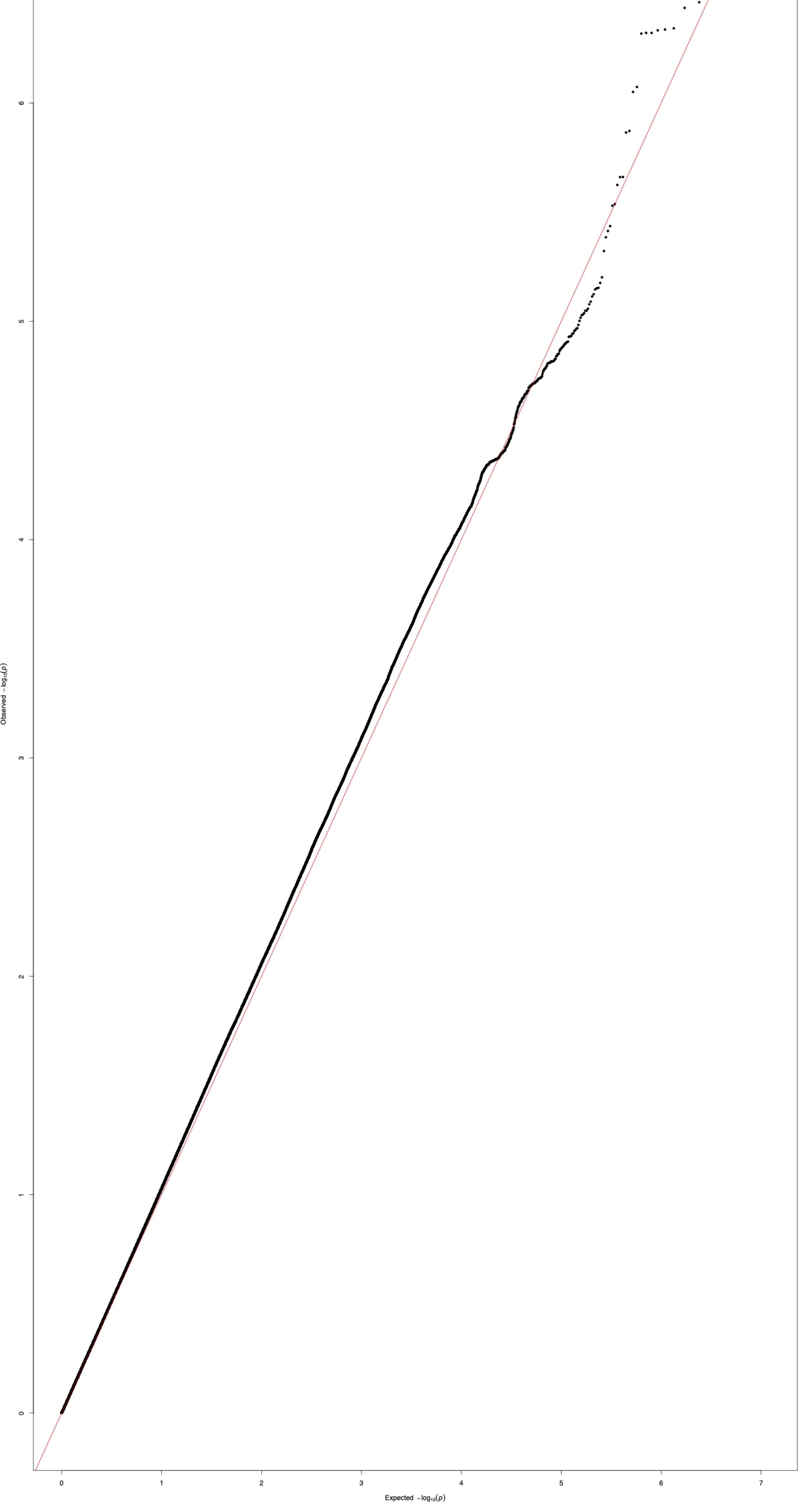
